# Identifying COVID-19 peaks using early warning signals

**DOI:** 10.1101/2025.03.03.25323224

**Authors:** Joshua Looker, Kat S Rock, Louise Dyson

## Abstract

The SARS-CoV-2 (COVID-19) pandemic has had catastrophic effects on public health and economies. Around the world, many countries employed modelling efforts to help guide pharmaceutical and non-pharmaceutical measures designed to reduce the spread of the virus. Modelling efforts for future pandemics could use the theory of early warning signals (EWS), which aims to predict ‘critical transitions’ in complex dynamical systems. In infectious disease systems, such transitions correspond to (re-)emergence, peaks and troughs in infections which can be indirectly observed through the reported case data. There is increasing evidence that including EWS in modelling can help improve responses to upcoming increases or decreases in case reporting. Here, we present both theoretical and data-driven analyses of the suitability of EWS to predict epidemic transitions in reported case data. We derive analytical statistics for a variety of infectious disease models and show, through stochastic simulations of different modelling scenarios, the applicability of EWS in such contexts. Using the COVID-19 reported case dataset from the United Kingdom, we demonstrate the performance of a range of temporal and spatial statistics to anticipate transitions in the case data. Finally, we also investigate the applicability of using EWS analysis of hospitalisation data to anticipate transitions in the corresponding case data. Together, our findings indicate that EWS analysis could be a vital addition to future modelling analysis for real-world infection data.

**Author summary:** The application of early warning signals (EWS), which uses statistics to predict changes in real-world data, has increasingly shown promise in successfully anticipating disease (re-)emergence or elimination of infectious diseases. We extend the existing literature through a theoretical study of EWS indicators under a variety of modelling scenarios that could have occurred during the COVID-19 pandemic (and in future pandemics). We show that EWS calculated on hospitalisation data can also accurately anticipate changes in the cases of related regions whilst also showing that spatio-temporal trends in EWS can be used to identify regions of concern.

## Introduction

The end of 2019 saw the beginning of the SARS-CoV-2 (COVID-19) pandemic in Wuhan, China. A highly transmissible pathogen with the possibility of causing long-lasting and severe complications, COVID-19 was quickly evaluated by the World Health Organization to be a public health emergency of international concern and by 11th March 2020, following case reporting in 114 countries, a pandemic [1]. Many countries, including the United Kingdom (UK), enacted non-pharmaceutical interventions (NPIs), alongside later mass vaccine rollouts, aimed at reducing transmission [2]. Across the globe, countries employed various NPI measures including the closure of schools and workplaces, social distancing, gathering size limits, isolation and quarantine routines, and travel restrictions [3–5]. In the UK, the government took similar measures during the 2020–2021 period. These included national lockdowns (e.g. in November 2020), gathering size restrictions (April 2021) and closures of workplaces and schools (October 2020). Naturally, the timeliness of these interventions directly influenced their effectiveness in decreasing the case numbers. Furthermore, to balance pandemic impacts on public health and on the economy, the UK government also undertook programmes to encourage resocialisation and increase spending (such as ‘Eat Out to Help Out’ in August 2020) and removed previously imposed restrictions, thus likely increasing virus spread [6].

Many governments also used mathematical modelling to predict the dynamics of COVID-19 with and without control measures, to make better-informed decisions on intervention policies. Modellers used a mixture of deterministic and stochastic methods to model the contagion dynamics and related statistics, such as reported cases, hospitalisations, and the effective reproduction number. Perfect mechanistic models could in theory be used to predict the future re-emergence of COVID-19 waves, but are in practice limited by the effects of data quality and human responses (such as the inability to accurately predict the implementation of intervention measures and public response to them). Model-independent methods such as early warning signals (EWS) have shown promise in anticipating critical transitions (in epidemics corresponding to peaks, troughs and (re-)emergence in the reported case time series) irrespective of the data quality and statistical noise [7–9].

In epidemic systems, modellers are interested in identifying the critical transition corresponding to a disease trending towards elimination, thus allowing for the end of control measures. Also of interest is the transmission potential of novel or re-emergent diseases. This potential can be quantified by the basic reproduction number, *R*_0_, the average number of secondary infections caused by a single infectious individual in a fully susceptible population [10]. When *R*_0_ *<* 1, the disease-free equilibrium is stable and the disease requires imported infections for sustained transmission. However, with *R*_0_ *>* 1 the disease-free equilibrium is instead unstable and a stable endemic equilibrium may be reached, provided there is sufficient replenishment of susceptible individuals. The critical transition between outbreak and elimination therefore occurs when *R*_0_ = 1 [7, 11–13].

The theory of critical slowing down (CSD) states that on the approach to these transitions, the system takes longer to return to its equilibrium after small external perturbations, causing predictable changes in the statistics of the time series (such as an increase in variance and the autocorrelation). These expected changes then provide advanced warning (early warning signals) of the upcoming transition, which have been successfully applied in the literature [7, 12, 14–21], with the aim of taking appropriate action to mitigate the resultant effects. CSD-based EWS have also been shown to predict non-stationary or non-linear periods (for epidemics, when there is growth or reduction in infections) that do not necessarily correspond to critical transitions —this is also of benefit in predicting further infection waves which are not generally caused by critical transitions [13, 16, 22, 23].

In reality, infection- or vaccination-conferred immunity and intervention measures reduce the susceptible pool available or the transmission rate, thus changing the effective reproduction number (*R*_*t*_) [10]. For scenarios where the susceptible pool has been depleted, we could define this as *R*_*t*_ = *R*_0_*s ≤ R*_0_, where *s* is the proportion of the population who are susceptible. It should be noted that *R*_*t*_ can also be further modified by interventions aimed at reducing infectious contacts (such as NPIs), though these will also affect *R*_0_. When *R*_*t*_ *>* 1, multiple secondary cases are likely to arise from every infection and the number of infections is likely to increase, but with *R*_*t*_ *<* 1 there is, on average, less than one secondary case from every infection and the disease will eventually die-out. Along the trajectory of an endemic dynamical disease system, this *R*_*t*_ = 1 transition can occur during (re-)emergence, at the peak of (re-)infection waves and on approach to elimination [16]. Although *R*_*t*_ = 1 has been referred to as a critical transition in the literature (and also due to confusion between *R*_0_ and *R*_*t*_) [16, 22, 24–26], it is not always a bifurcation (as it is also a function of the system state) and does not indicate a change in the stabilities or values of the system equilibria. Nevertheless, this point does represent qualitatively different behaviour in the dynamics of the epidemic system and it has repeatedly been anticipated through EWS analysis based on CSD indicators [12, 16, 22, 24, 25].

Indeed, CSD and other EWS analyses have been investigated in the context of the COVID-19 pandemic (primarily using country-level incidence data) with varying levels of success [16, 22, 24, 27, 28]. These studies investigated whether EWS could be used to predict both re-emergence and waning. The fast dynamics of the pandemic, however, caused difficulties in reliably predicting these changes and accurately estimating the long-term trend in the observed cases (required to calculate the stochastic perturbations) [22, 24, 28]. Furthermore, recent results in the literature suggest that EWS show spatio-temporal effects that could be more noticeable than only considering the temporal domain [26, 27]. Spatial detrending methods (using the average of multiple populations that are anticipated to have similar characteristics) have also been found to give better EWS [29], suggesting that having high spatial granularity in COVID-19 case data could lead to better early warning signal performance. In the context of the COVID-19 pandemic, spatially-fine-grained datasets have been successfully used to anticipate local outbreaks through an increase in the local network entropy [27].

The UK COVID-19 data has a far finer level of granularity and more consistent reporting than most previous outbreaks. Publicly-available daily case data was published for 315 regions/cities/smaller areas called Lower Tier Local Authorities (LTLAs), with other data (including hospitalisations) also published for the seven National Health Service (NHS) regions. LTLAs are the smallest statistical area for which case data is publicly available in the UK, and correspond to the local governments of associated metropolitan, non-metropolitan, unitary authorities and London Boroughs. Populations range from 40,476 in Rutland to 1,140,525 in Birmingham and areas from approximately 12km^2^ in Kensington and Chelsea to 5000km^2^ in Northumberland. The seven NHS regions are geographic divisions of England representing the seven different NHS regional teams that each serve different LTLAs and oversee all NHS organisations within their region. Thus, this infection dataset provides a unique opportunity to investigate whether changes in temporal (such as the variance) and spatial (such as the spatial skew) statistics reliably anticipate upcoming critical transitions in a real-world disease system.

In this paper, we investigate whether using EWS analysis alongside other modelling efforts could have impacted decisions made in the June 2020 to December 2021 period of the COVID-19 pandemic in the UK. We first consider, through simulation and theory, the ability of EWS to anticipate the emergence of new variants and the approach of epidemic peaks. Since previous studies in the literature investigate whether CSD occurs before the (re-)emergence and suppression transitions of *R*_*t*_ = 1, we were motivated to consider whether EWS analysis can also anticipate the equivalent change in *R*_*t*_ at the peak of infection waves. Using the publicly-available UK Health Security Agency (UKHSA) reported case and hospitalisation data, we consider the ability of various temporal and spatial statistics to anticipate these critical transitions. Finally, we employ the “2-*σ* detection method” [7] to quantify how early these transitions could be anticipated. This allows us to explore whether this advanced warning would have been enough to have changed policy decisions made during the selected pandemic period.

## Materials and methods

Here we summarise the methods used, beginning by deriving the expected behaviour of various EWS in infectious disease models on the approach to an epidemic transition, and detailing the simulation methods used. We then describe the dataset and pre-processing required, along with a summary of the course of the UK pandemic and how EWS were applied to the data, including investigation of the timing of EWS.

To calculate the EWS and analyse the results we used Python 3.11 with packages: Numpy (version 1.26.4), Matplotlib (version 3.84), GeoPandas (version 0.14.4), statsmodels (version 0.14.1), epyestim (version 0.1) and Pandas (version 2.2.1). A repository containing the data and code used to conduct this study can be found at https://github.com/joshlooks/covidews.

### Early warning signals: the theory

This section outlines the mathematical theory behind EWS upon the approach of an epidemic transition. We explore possible mathematical models for the (re-)emergence and peaks of COVID-19 waves (due to the easing of restrictions or mutation of new variants) and the effects of NPIs. Analytical EWS results are derived from the Fokker-Planck equations (FPEs) and corresponding stochastic differential equations (SDEs) for the stochastic deviations from the mean for two common disease models. We selected the susceptible-infectious-susceptible (SIS) model as a baseline to match with the existing literature. The susceptible-exposed-infectious-recovered model (SEIR) is also analysed as it is commonly used to model COVID-19 infection and, to the best of the authors’ knowledge, has not yet been theoretically considered under an EWS framework although numerical analyses have been conducted [25, 26]. We use extensive Gillespie simulations to explore EWS results under a variety of situations (re-emergence, NPI introductions, change of social behaviours) present during the COVID-19 pandemic. Finally, we also investigate the impacts of over-dispersion and reporting probability in case reporting on prediction power.

#### Analytical early warning signals

On the approach to an epidemic transition, the theory of critical slowing down states that there will be an increase in the return time of stochastic fluctuations. This can be observed as predictable trends in the statistics of fluctuations from the long-term trend in the infectious time series. To develop an understanding of the possible behaviour of such statistics, we derived the SDEs and corresponding FPEs for the stochastic deviations from the long-run averages of the infectious compartment in two commonly used disease models: the SIS and SEIR models. Finally, we solved for the moments of the FPEs to analyse their expected behaviour on the approach to a critical transition.

We began with a simple SIS model of disease (re)-emergence due to the introduction of a new variant (with full derivations in S1 Text) where we considered the effective contact rate (*β*) between susceptible (*S*) and infectious (*I*) individuals as the control parameter. This allowed for similar analysis to be considered for the introduction or easing of NPIs which also impact *β*. Assuming a constant population size of *N* = *S* + *I*, the corresponding system is then one-dimensional with the ordinary differential equation (ODE) for *I* as:

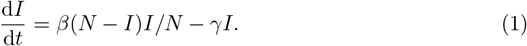

where *γ* is the recovery rate of infectious individuals. We took the approach in Schnoerr *et al*. [30] in using the system size expansion and linear noise approximation [12, 30, 31] to derive the FPE for the probability distribution (Π(***ζ***, *t*)) of the stochastic fluctuations in the system:

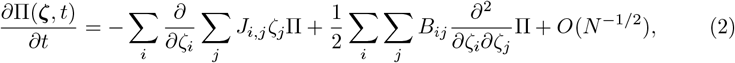

where *J* is the Jacobian of the system (with respect to the average proportion of individuals in each compartment), the subscripts *i, j* represent each compartment and transition in the model, respectively, *ζ*_*i*_ is the stochastic fluctuations from the average proportion of individuals in compartment *i* and *B* is the noise-covariance matrix of the system.

Returning to the SIS system, the FPE and corresponding SDE can then be derived (S1 Text). Finally, we could obtain the differential equation for the analytical variance (*⟨ζ*^2^*⟩*) as in Southall *et al*. [32]:

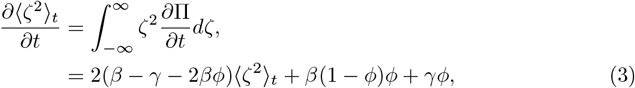

where *ϕ* = *I/N* . Previous numerical results for this equation show an increasing variance on approach to critical transitions (as expected from CSD theory) [12, 29, 32].

We wish to understand EWS in the SEIR model with demography (Fig 1), as similar systems have been used successfully to model COVID-19 [33, 34]. We present a fuller analysis of this system as it has not previously been studied using the linear noise approximation in the literature.

**Fig 1.**
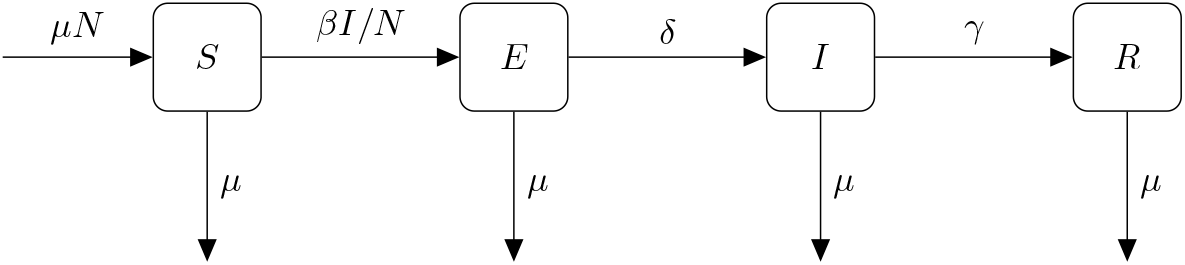
SEIR compartmental diagram showing the infection, recovery, birth/death and symptom onset processes. *β* is the effective contact rate between susceptible and infectious individuals, *γ* the rate of recovery of infectious individuals and the expected latent period is 1*/δ*. The birth and death rates (*µ*) are set equal such that the population size is constant.

The corresponding model equations are:

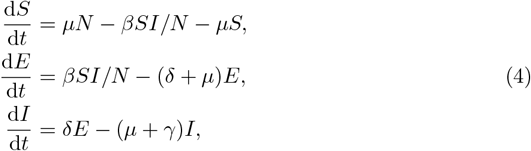

where 1*/δ* is the expected latent period and *µ* is the natural death/birth rate. We note that by equating the birth and death rates, the average population size is constant such that *S* + *E* + *I* + *R* = *N* and the ODE system is three-dimensional. The SEIR model also has two fixed points, a disease-free equilibrium (*S*^*\**^ = *N* ) and an endemic equilibrium 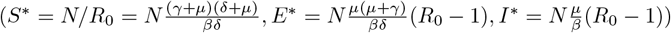. For the deterministic system, or in a sufficiently large population, it is well known that the endemic equilibrium is stable for *R*_0_ *>* 1 (and unstable for *R*_0_ *<* 1), while the disease-free equilibrium is unstable for *R*_0_ *>* 1 (and stable for *R*_0_ *<* 1) [10].

Next, we used the linear noise approximation to express the number of susceptible, exposed and infectious individuals (*S, E, I*) in terms of the expected proportion of the population who are susceptible, exposed and infectious (*ϕ*_1_, *ϕ*_2_, *ϕ*_3_, respectively) and the deviations from these mean-field dynamics (*ζ*_1_, *ζ*_2_, *ζ*_3_) as:

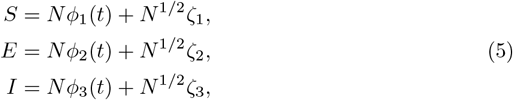

where again *ϕ*_*i*_, and *ζ*_*i*_ are the expected proportion and stochastic deviations. To first order (in *N* ), Eqn 4 is then given as:

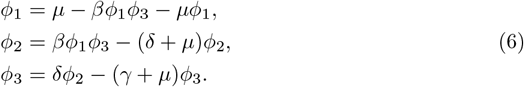

Finally, we needed to find the Jacobian and noise-covariance matrices to express the Fokker-Planck equation of the system (*ζ*_1_, *ζ*_2_, *ζ*_3_). The Jacobian is:

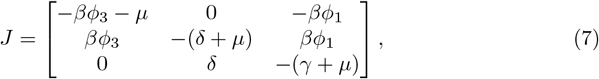

and the noise-covariance matrix is:

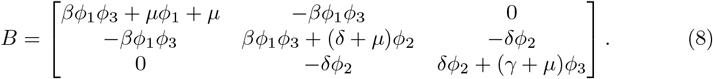

Although there have been many temporal EWS proposed in the literature associated with transitions (trends in summary statistics such as the variance, autocorrelation, skewness and kurtosis), the variance has consistently performed best in terms of predictive power (generally measured using the area under the curve) [7, 8, 17, 26]. To calculate the variance, we took the approach used in Southall *et al*. [32] by numerically solving the Lyapunov equation for the evolution of the covariance matrix Θ_ij_ = ⟨ζ_i_ζ_j_⟩ − ⟨ζ_i_⟩⟨ζ_j_⟩:

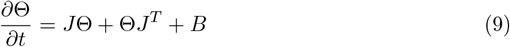

where the variance of the infectious fluctuations is given by Θ_33_. We note that this (linear noise) assumption then also leads to the number of individuals in each compartment (*X* = (*S, E, I*)) following a multivariate normal distribution given by:

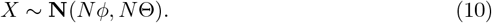

We have successfully derived the distribution of stochastic fluctuations in the SEIR model. This allowed us to next analyse the expected behaviour of EWS on approach to epidemic transitions.

#### Gillespie simulations

We next investigated the behaviour of EWS for the SEIR model for four scenarios: with fixed effective contact rate; with an increasing effective contact rate (due to the relaxing of NPIs/change in behaviour/introduction of a new variant); with continuous decrease in effective contact rate (due to NPIs); and with a step-wise decrease in effective contact rate (due to an immediate NPI introduction) to analyse whether epidemic transitions can be predicted or bifurcation delays can be observed (Fig 2).

**Fig 2.**
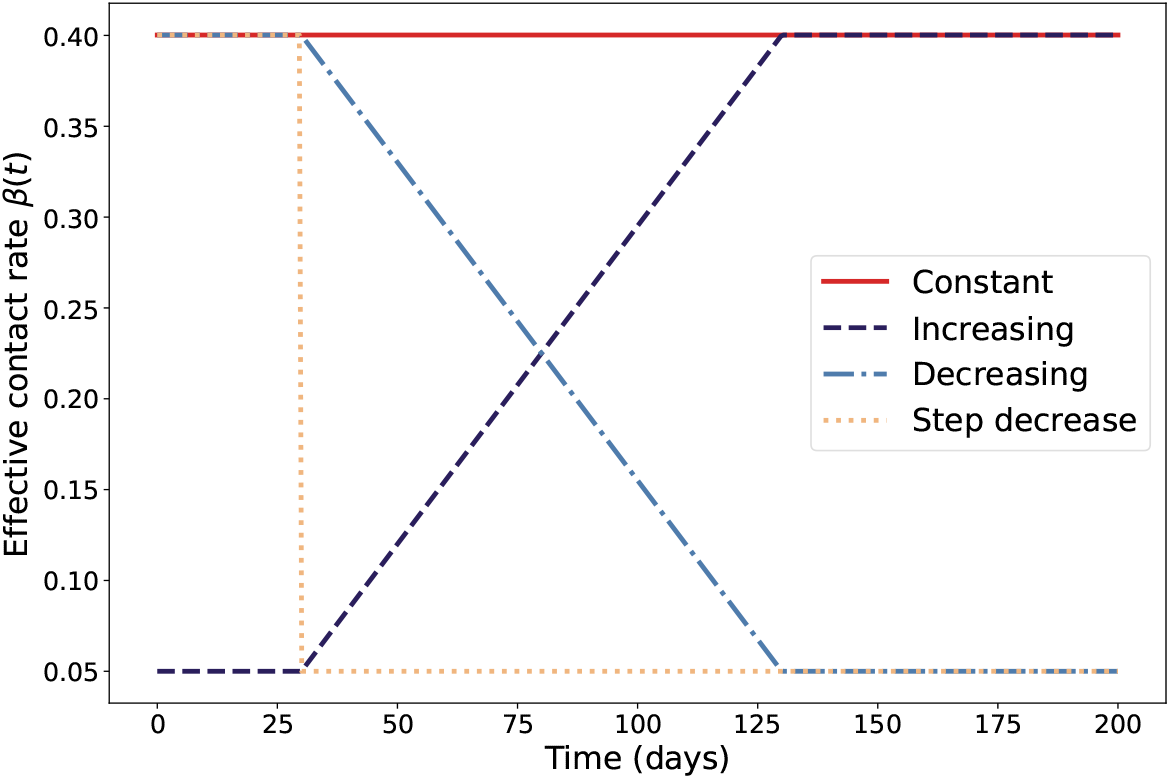
Effective contact rate over time for the different modelling scenarios considered: fixed, continuously increasing, continuously decreasing and with a step-decrease.

The model parameters, shown in Table 1, are informed by previous SEIR-based modelling of the wild-type and Alpha variants of COVID-19, and biologically relevant distributions (such as the latency period) [33–38], and differ in the form of *β*(*t*) (which we currently consider to have a different time-dependent form depending on the modelling scenario). The basic reproductive number is assumed to be *R*_0_ = 4 for the Alpha-variant, and *R*_0_ = 3.1 for the wild-type variant, when setting the base effective contact rate (with the corresponding base effective contact rates similar in value to those used in [33, 34]). Note that these simulations are not designed to perfectly capture the spread of the two variants considered within the UK but to test the identifiability of EWS for a wild-type-like and an Alpha-like contagion undergoing SEIR dynamics.

**Table 1.** Parameters and values for each set of simulations run. The parameters are chosen to give similar contagion dynamics to both the wild-type and Alpha variants of COVID-19 in populations of the same magnitude as the three LTLAs considered in the UKHSA reported case results (Maidstone, York and Camden).

| Parameter | Biological Interpretation | Value |
| --- | --- | --- |
| $\beta_0$ | Effective contact rate (baseline) | 0.4002, 0.3102 per day (Alpha-like, wild-type-like) [35, 40, 41] |
| $\mu$ | Natural mortality/birth rate | $1/(80 * 365)$ per day (both pathogens) |
| $1/\delta$ | Latent period | 5 days (both pathogens) [33, 34] |
| $1/\gamma$ | Infectious period | 10 days (both pathogens) [33, 34] |
| $\beta_{low}$ | Effective contact rate (lower due to NPIs or similar) | 0.05 per day (both pathogens) |
| $p$ | Change in effective contact rate (due to NPIS or similar) | $(\beta_0 - \beta_{low})/100$ per day (both pathogens) |
| $\tau$ | Day $\beta(t)$ starts to change (for scenario two and four) | 30 days (both pathogens) |

We also note that both dynamic bifurcations (*R*_*t*_ = 1) corresponding to the peak in each infection wave and the more standard equilibrium bifurcation (*R*_0_ = 1) are present in this set of modelling scenarios. This allowed us to investigate whether there are any differences in the slowing down observed on approach to the different types of transitions seen in real-world disease data. For the baseline of a fixed effective contact rate, there is only a dynamic transition, as *R*_0_ *>* 1 throughout the simulations. The simulations with linearly decreasing *β*(*t*) include both types of transition, with the *R*_0_ = 1 equilibrium bifurcation occurring almost immediately after the *R*_*t*_ = 1 transition. Increasing *β*(*t*) simulations also have an *R*_0_ = 1 bifurcation, which also corresponds to the approximate point where *R*_*t*_ approaches one from below. This is then followed by another epidemic transition at the peak. We expected to see dynamic instability on approach to the peak in the first two sets of simulations, and on approach to both transitions in the increasing *β*(*t*) scenario. However, due to the instant forcing through the bifurcation point in the step-decrease scenario, we do not expect a rise in dynamic instability on approach to this point. Nevertheless, this scenario allowed us to investigate whether there are predictable signals that would allow modellers to confirm the efficacy of a lockdown (or similar NPI that forces a step-change in contact rate) in suppressing transmission.

We used the Gillespie method [39] to run 10,000 simulations of each of the four modelling scenarios, for the two different COVID-19-like variants, totalling eight sets of simulations. Event and transition rates for the four scenarios are given in Table 2. Each simulation had a population size of *N* = 100, 000, an initial infectious population of 10 individuals, with the remaining 99,990 individuals being susceptible and a maximum simulation time of 200 days. The population size was chosen to be of the same magnitude as the three LTLAs analysed in depth.

**Table 2.**
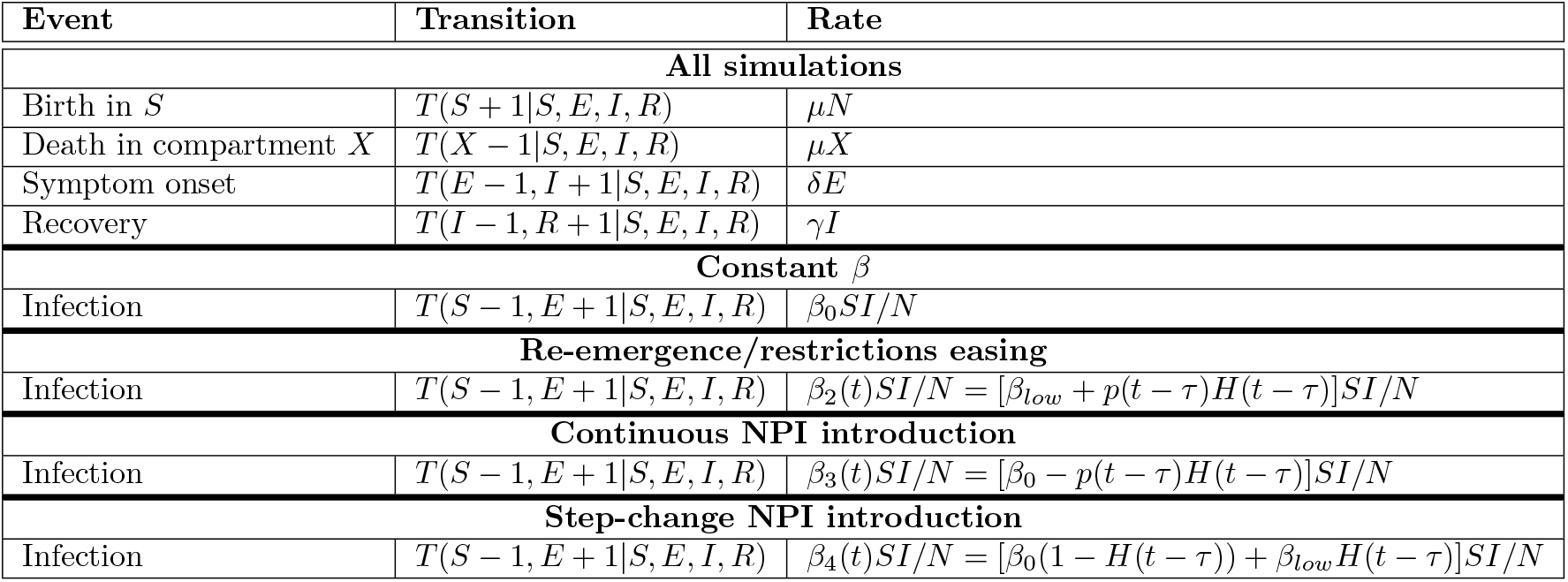
Events and transition rates for the four sets of Gillespie simulations run. Here *T* (*Y* |*X*) indicates a transition from a system state of *X* to a system state of *Y* . The first set has a constant effective contact rate. The second models the re-emergence of infection due to the easing of restrictions or the introduction of a new variant and has *β*_2_(*t*) = *β*_*low*_ + *p*(*t* − *τ* )*H*(*t* − *τ* ). The third and fourth model the introduction of NPIs continuously (*β*_3_(*t*) = *β*_0_ − *p*(*t* − *τ* )*H*(*t* − *τ* )) and as a step-change (*β*_4_(*t*) = *β*_0_(1 − *H*(*t* − *τ* )) + *β*_*low*_*H*(*t* − *τ* )) respectively.

We analysed the simulation results in a similar way to the COVID-19 data (which can be seen in the Simulation results section), which will be explained in the following subsections. The main difference is that when using the Gillespie simulations the prevalence time series was detrended by subtracting the time-dependent average of all realisations (of that modelling scenario). Normalised statistics of the detrended time series were then analysed using the 2-*σ* algorithm (see the Detecting an epidemic transition section) to determine when the upcoming transition was detected. The estimated location of the epidemic transitions, and critical transitions where appropriate, was found by taking the average time *R*_*t*_ = 1 from the simulations in each scenario.

After investigating whether the linear noise approximation and standard EWS theory can be applied to the prevalence of infectious individuals, in a best-case scenario of “perfect information” we also analysed the effects of errors in case reporting on the ability to anticipate the epidemic transitions. We took a similar approach to O’Dea and

Drake [42], and Brett *et al*. [18] in modelling the reported cases by applying a negative binomial reporting error to the incidence of infections in the alpha-like simulations (which we took as the *E → I* transition in the model. The aggregation period for cases was taken as one day to match the COVID-19 reporting period, and we varied both the reporting probability, *ρ*, and dispersion, *ξ*. The probability distribution for the number of cases at time *t, K*_*t*_, is then given as

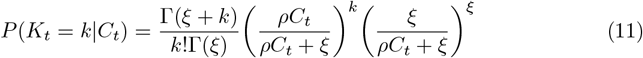

where *C*_*t*_ is the number of *E → I* transitions in the aggregation period. As noted in Brett *et al*. [18], the mean number of reported cases is then *ρC*_*t*_ and the variance is given as 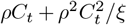, where an increase in *ξ* reduces the overdispersion in the data such that for high *ξ* the distribution is approximately Poisson (with equal mean and variance).

We selected *ρ* = 0.8, 0.6 as two possible reporting probabilities, and dispersion parameters of *ξ* = 1, 10. Setting *ξ* to any lower order of magnitude, for example *ξ* = 0.1, leads to biologically-infeasible case statistics, with more cases reported at peak than individuals in the population.

### Early warning signals: the data and methods

In this section, we summarise the pre-processing conducted on the UKHSA dataset to improve the forecasting skill of the EWS considered and to standardise the dataset into an interpretable form. We also describe the EWS considered in the study and how they are numerically calculated.

#### UK COVID-19 data: incidence and hospitalisations

Although COVID-19 infection data is still being published today, we focused on the period from June 2020 to December 2021 as it contained several important events that impacted the infection dynamics, as shown in Fig 3. The changes in *R*_*t*_ caused by these events lead to smaller (re-)infection waves that could possibly have been anticipated by EWS analysis. This period saw the introduction of the ‘Eat Out to Help Out’ campaign in August 2020 by the UK government to support the hospitality sector [6]. Coupled with the easing of social restrictions in June and July, these changes led to the start of a COVID-19 wave in September 2020 [6]. After the introduction of additional measures (including enforced work from home) in September and October, there was another period of non-stationarity in the reported case time series. Finally, the dominance of the Alpha-variant in the December 2020 to January 2021 time period (S6 Fig) led to a much larger COVID-19 wave in the 2020 Christmas and New Year period. We note that a vaccination campaign against COVID-19 in the UK began in the January–February period of 2021, with the majority of the UK receiving a first dose within the first few months. Previous studies have shown that second doses are required for effective reduction in transmissibility and disease severity (especially for Alpha, Delta and Omicron variants). Since less than 65% of the population received a second dose, we therefore do not expect the vaccination to noticeably reduce the last transition that we consider (in July 2021). Therefore the vaccine was not an indicator of a critical transition [43–45]. We also note that the data between June and December 2021 shows periods of non-stationarity likely caused by the emergence and dominance of the Delta variant.

**Fig 3.**
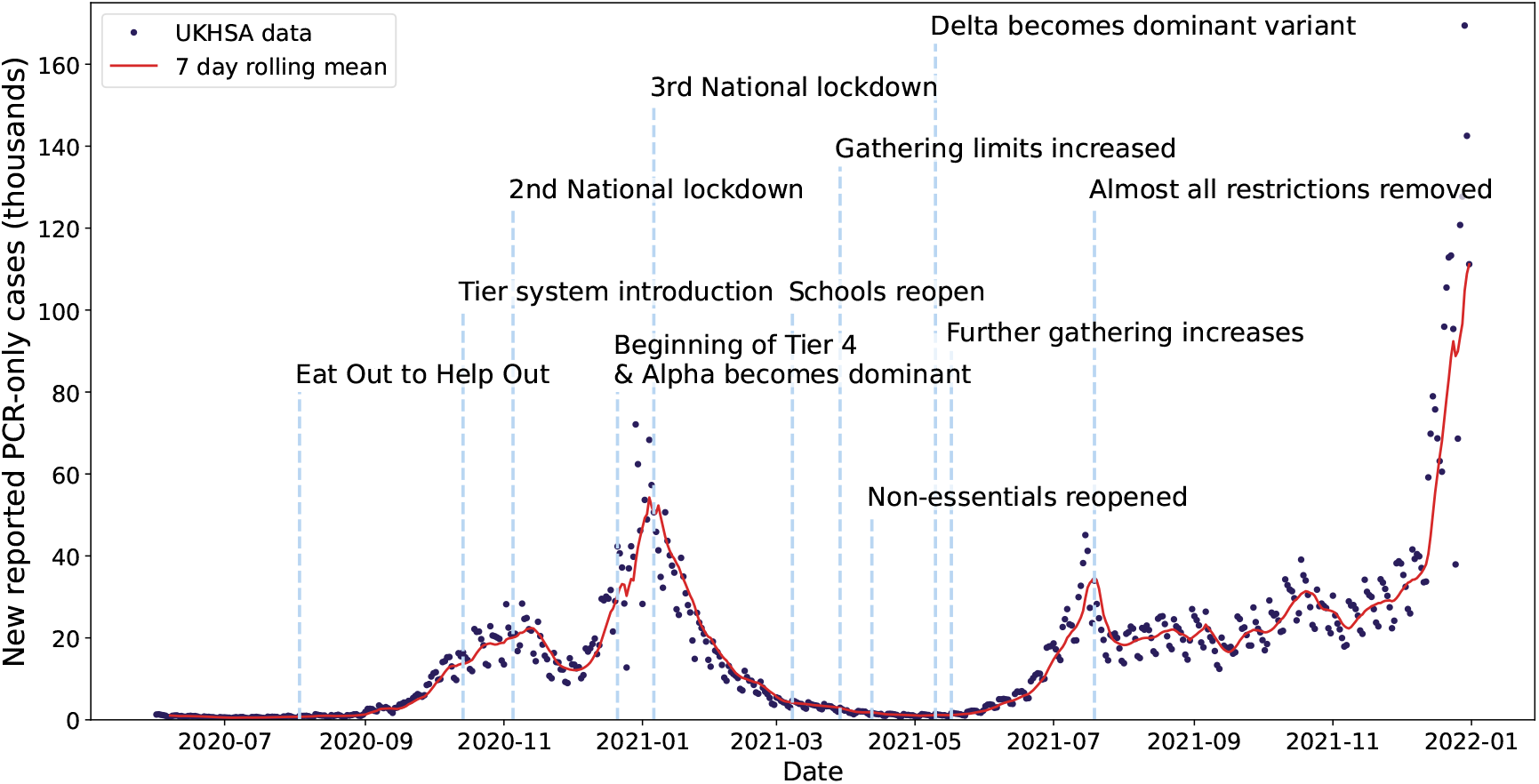
Timeline of the COVID-19 pandemic in the United Kingdom between June 2020 and December 2021. Daily reported polymerase chain reaction-only case data is displayed as dark dots, and the red line shows the rolling seven-day average. Key NPIs or changes in restrictions are noted: Eat Out to Help Out [6], Tier system introduction [46], 2nd National lockdown [46], Beginning of Tier 4 & Alpha becomes dominant [47], 3rd National lockdown [46], Schools reopen [48], Gathering limits increased [49], Non-essentials reopened [49], Delta becomes dominant variant [50], Further gathering increases [46], Almost all restrictions removed [46].

Two types of COVID-19 tests were commonly used in the UK: polymerase chain reaction-only (PCR) tests, which test for the presence of viral ribonucleic acid, and lateral flow device (LFD) tests, which test for viral antigens. The potential usable datasets are then: PCR-only, PCR-confirmed LFD, and LFD only (collected by UKHSA for their, now decommissioned, COVID-19 dashboard [51]). As during the time period considered almost all reported tests were PCR tests, we therefore chose to use the PCR-only dataset. There are also two potential routes for PCR test: ‘Pillar 1’ (healthcare-related testing) or ‘Pillar 2’ (mass community testing). The dataset used is primarily composed of Pillar 2 testing.

To smooth the data, we aggregated the daily incidence of newly reported PCR-only cases to seven-day rolling sums. With a high daily variance in testing rates, this reduces the external noise in the resultant reported case time series. Using this summation also removes the ‘weekend’ effect seen (whereby testing and reporting rates were lower during the weekend [16, 22]). The daily reported case time series was analysed at the level of the 315 LTLAs (the smallest statistical area considered by the Office for National Statistics) within the UK, with the EWS calculated for each LTLA individually.

We focused on three LTLAs in particular for individual reporting: Maidstone, York and Camden. These LTLAs are spread across the UK with Maidstone in the South East, Camden in London, and York in the North East. With the Alpha-variant hypothesised to have originated in East London or Kent [52], we also expected the transition times to differ between these LTLAs, particularly in late 2020. Further, the LTLAs selected have different population sizes, so the impacts of population size on EWS, and the level of noise in the statistics, could also be examined (Maidstone with 173,132, Camden with 279,516 and York with 211,012 inhabitants according to the 2021 census [53]).

In addition to the PCR-only case reporting data, we also considered temporal statistics calculated on both the hospitalisation incidence and occupancy time series recorded by UKHSA [51]. These were collected for each of the seven NHS regions in the UK and are also smoothed by aggregating to seven-day rolling sums.

For each of the aggregated time series considered, the values are normalised by dividing by the population size of the corresponding region (LTLA or NHS region). This allows for the comparison of values across the different areas and for spatial/metapopulation detrending to be conducted (see the Calculating early warning signals section).

#### Calculating early warning signals

This section outlines the numerical calculation of the statistics considered (both temporal and spatial) in the EWS analysis of COVID-19 reported case time series, including the window sizes used in the estimations. We also explain the detrending techniques used to separate stochastic fluctuations in the time series from the long-term trend.

The theory of EWS identifies expected trends in the statistics of stochastic fluctuations away from the long-term trend of the time series. We therefore took a metapopulation approach to detrend the smoothed time series (shown to be more accurate than purely temporal methods such as Gaussian or rolling-window detrending, [27, 29]). For the LTLA-reported case time series, the average value for the corresponding NHS region was subtracted from each LTLA within that region. For the NHS regional hospitalisation incidence and hospitalised occupancy time series, we instead subtracted the national average.

After aggregating, normalising by the population size and detrending the time series, we calculated EWS on the resulting residuals. For each of the temporal statistics considered, we took an online approach: calculating the statistics on each day using only previously occurring data, mirroring how EWS would be applied in a real-world setting. The statistics were calculated using a back window of seven days, for those calculated on case incidence and hospital occupancy, or fourteen days, for those calculated on hospitalisation incidence. These window sizes were set using a sensitivity analysis to account for the differing noise levels, although a full analysis was also conducted using window sizes of 10, 14 and 30 days for the case incidence in the Supporting Information. For temporal statistics, we considered both the variance and first difference in variance for each of the LTLAs. Other potential statistics were calculated but did not lead to robust EWS of upcoming transitions (S7 Fig) [22, 28].

The calculations were done using the Python package ‘Pandas’, which uses unbiased population estimates of statistics (for example the variance and skew). The variance (*V* (*t*)) and first difference (*FD*(*t*)) were calculated as:

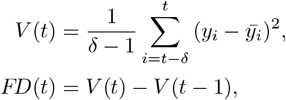

where *δ ∈ {* 7, 14*}* is the window size used and *y*_*i*_ and 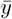 are the residual and mean at time point *i*, respectively.

The two types of spatial statistics considered are spatial variance and skew, which have previously shown promise in anticipating critical transitions [29, 54–57]. These were calculated at the NHS regional level by considering the variance and skew across the residuals of all LTLAs within an NHS region. Explicitly, the spatial variance (*SV* ) and spatial skew (*SS*) were calculated as

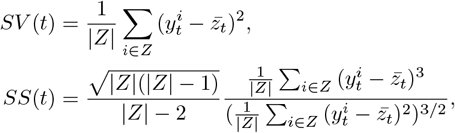

where 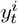 is the residual of LTLA *i* at time point *t*, 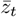 is the average of all LTLAs in the NHS region *z* at time point *t, Z* is the set of all LTLAs in NHS region *z*.

We also note that the normalised versions of these statistics were calculated to be used alongside the 2-*σ* method to detect outlying values (explained in the Detecting an epidemic transition section). To normalise the statistics, we subtracted the expanding mean and then divided by the expanding standard deviation (Fig 4 bottom left). The rolling mean and rolling standard deviation were also tested but did not affect the results.

**Fig 4.**
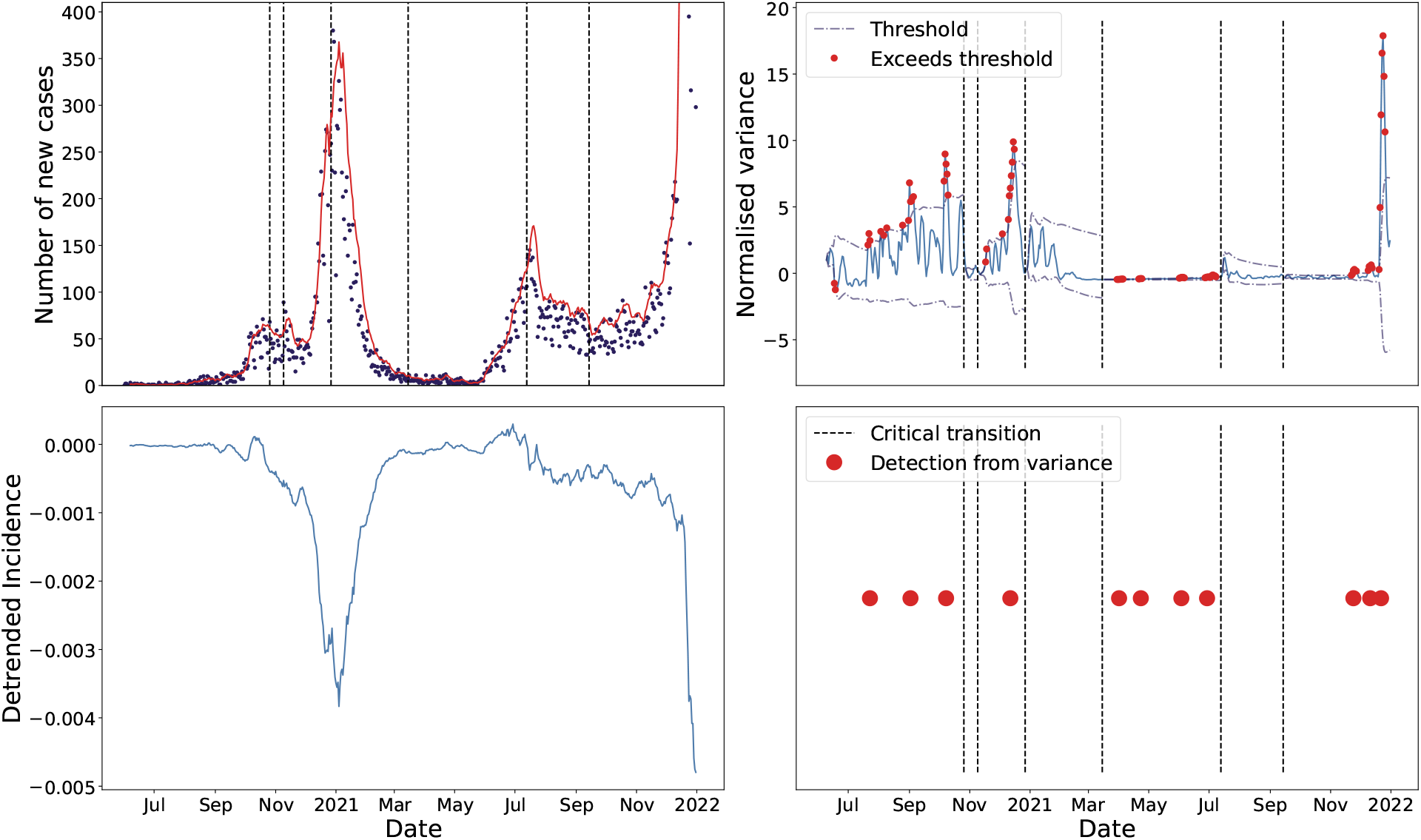
Full method used for detecting EWS applied to the COVID-19 reported case time series in Camden. Top left, number of reported PCR-only COVID-19 cases (black dots) with the seven-day rolling sum overlayed. The vertical dotted lines correspond to the approximate date that *R*_*t*_ = 1. Bottom left, the detrended residuals in the reported case time series with the average incidence of the corresponding NHS region used for detrending. Top right, the normalised variance in residuals, with the dashed lines displaying the 2-*σ* detection threshold, the red dots signifying exceedances of this threshold and the dotted vertical lines again showing the critical transitions (whereon the threshold is reset). Bottom right, the resultant time-of-detection timeline showing the time-of-detection for upcoming critical transitions as the red dots (with each corresponding to three consecutive threshold exceedances in the subplot above).

#### Detecting an epidemic transition

The 2-*σ* method from Drake [7] proposes that a normalised EWS is significant if its instantaneous value exceeds two long-run standard deviations on either side of its long-run mean value. Following [58], we took the time-of-detection for the upcoming epidemic transition to be when at least three consecutive time points cross the threshold. The 2-*σ* threshold is analogous to a 95% confidence interval for the statistic value. This method has also previously been used in the context of COVID-19 [22].

Similarly to Dablander *et al*. [16], we needed a method to determine where an epidemic transition (*R*_*t*_ = 1) occured. This allowed us to then analyse the effectiveness of different statistics in anticipating these transitions and also to refit the 2-*σ* method after each transition. Without refitting, time series statistics could be masked by previous transitions [16]. We made use of the method outlined in Huisman *et al*. [59] which builds on the method from Cori *et al*. [60]. The method smooths the reported case time series using local polynomial regression with a smoothing parameter that we set to 63 days at the LTLA level and 35 days at the NHS level (again set through a sensitivity analysis). This smoothed case time series was then used to estimate the number of infections and *R*_*t*_ using Bayesian inference techniques.

Although this may seem to combine online (*i*.*e*. real-time) learning and retrospective analysis, we note that in practice our method would only be used to predict the next upcoming epidemic transition. Changes in the statistics due to previous transitions can reduce the ability to anticipate future transitions. Therefore, once the transition has definitely occurred, the expanding mean and standard deviation calculations would be stopped and recalculated to anticipate the next transition (in the same manner that we conduct retrospectively). Explicitly, the calculations were run from the beginning of the time series until the first point that *R*_*t*_ crossed through one, from the last point that *R*_*t*_ until the next, or from the final *R*_*t*_ transition until the end of the time series. Finally, the EWS analysis method conducted (in terms of predicting the next upcoming transition) does not rely on fitting *R*_*t*_ and the fitting is only necessary to retrospectively evaluate how including EWS analysis may have impacted modelling efforts and pandemic response.

The complete method from reported case time series to time-of-detection is summarised in Fig 4.

## Results

In this section, we report the numerical results obtained from analysing the ability of EWS to reliably predict the re-emergence and die-out of subsequent COVID-19 waves between June 2020 and December 2021. We begin with theoretical results obtained from simulations designed to capture different behaviour from the pandemic. The data-driven analysis begins with the temporal statistics and time-of-detection results for incidence-based EWS at the LTLA level before reporting the corresponding analysis for the spatial statistics at the NHS region level. Finally, the EWS results calculated from hospitalisation data are also reported.

### Simulation results

The first section of results indicates the suitability of the linear noise approximation to the transient dynamics of an SEIR model and that early warning of upcoming peaks is detectable in both prevalence and case data. Simulation data from the sets of 10,000 Gillespie simulations run for the four different modelling scenarios considered, described in the Gillespie simulations section. All results were made using the Alpha-variant-like parameters (with results for wild-type-like parameters showing similar qualitative trends and presented in S2 Fig). To compare the Gillespie simulation outputs (at consistent time intervals), we aggregated the proportion of individuals in SEIR to daily counts from the start of each day. We could then calculate the infectious proportion over time for each modelling scenario considered (single SEIR wave, increasing effective contact rate, gradually decreasing effective contact rate, and step-decrease in effective contact rate: S2 Fig). We analysed how the ‘simulation’ variance in the proportion infected (variance between each simulation’s proportion infectious at each time point) and the time-of-detection distribution differ between scenarios (Fig 5). We see the variance increasing on approach to the epidemic peak in all scenarios tested (Fig 5, third column). There is a second spike in the variance after the epidemic transition for all scenarios bar the step-decrease in *β*, which could be used by modellers to anticipate an approach to endemicity. Under the linear noise approximation, the rate of change of the variance is dependent on the stability of the eigenvalues; i.e., the stability of the Jacobian controls the growth or decay of perturbations to the deterministic trajectory [61–63]. For the SEIR model with demography, a pair of negative, real eigenvalues near the disease-free equilibrium (or on its centre manifold) become complex conjugates with positive real part throughout the rest of the trajectory [10]. This leads to the double peak in variance due to the growth in oscillatory modes/perturbations as the real part of the eigenvalues goes through zero. Further, the variance in simulations, for both pathogens and all scenarios except increasing *β*, shows good concordance with the theoretical variance. For the ‘increasing *β*’ simulations, we observe a difference in magnitude between the theoretical and simulation-based variance. This is because we specifically selected simulations that did not fade out, thus increasing the correlation between the simulation results and reducing the variance between simulations (compared to theory). Nonetheless, the qualitative shape of the theory and simulation-based variances are the same which suggests that an increase in variance is still a good EWS of (re-)emergence.

**Fig 5.**
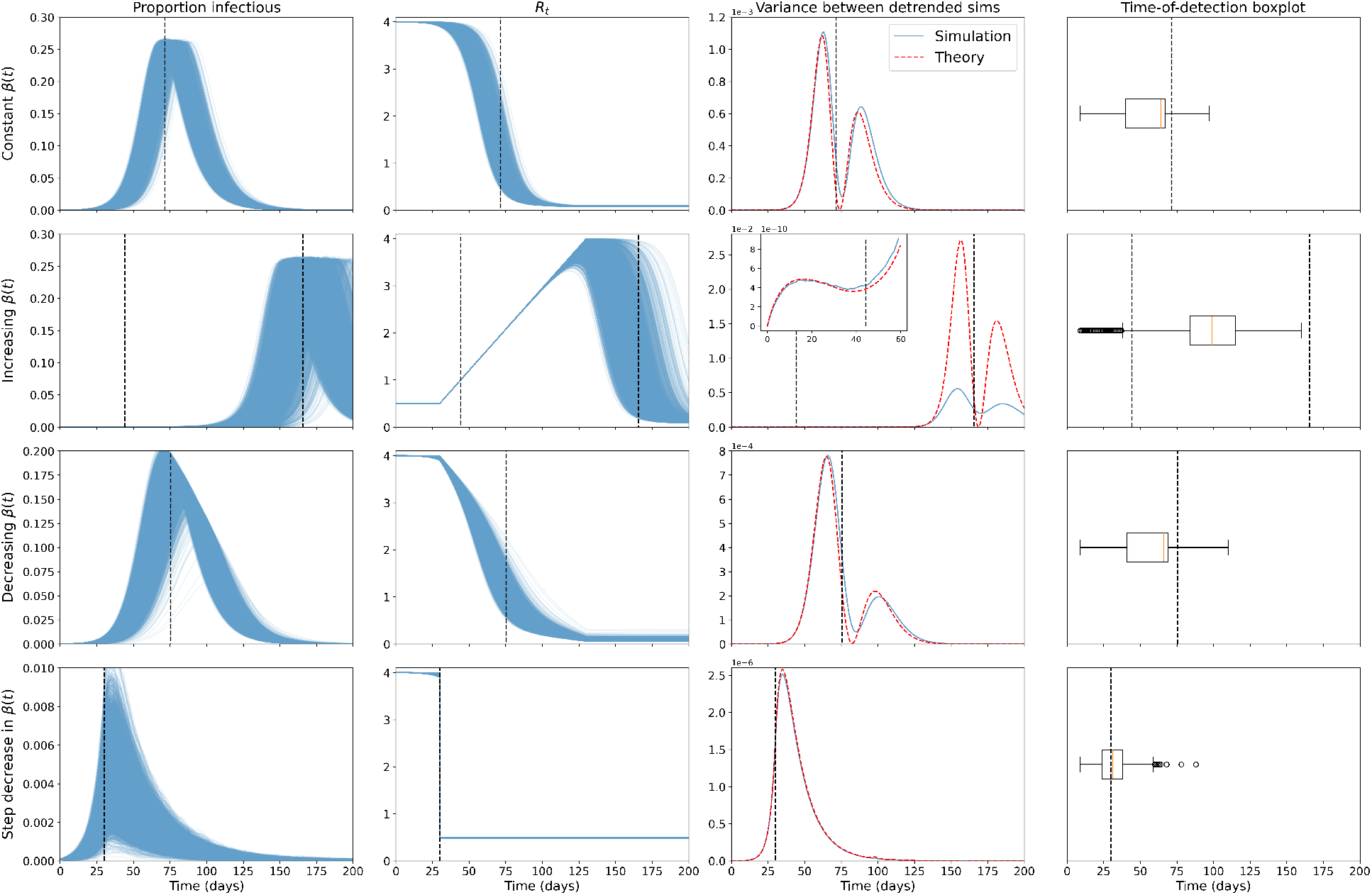
EWS results for the prevalence of the Alpha-like pathogen show successful anticipation of the epidemic transition. Columns show proportion infectious over time, effective reproduction number, variance between the mean-detrended simulations and time-of-detection distribution for the 10,000 simulations run for each of the four modelling scenarios (rows, constant *β*(*t*), increasing *β*(*t*), decreasing *β*(*t*) and a step-decrease in *β*(*t*)) for the Alpha-like pathogen.

In the increasing simulations (Fig 5, row three), we also see an increase in the variance on approach to the bifurcation of *R*_0_ = 1 at around 45 days. This increase is not monotonic, however, with the variance curve flattening around the bifurcation point.

The time-of-detection distribution (three consecutive 2-*σ* exceedances) in all simulations (for the Alpha-variant, Fig 5 fourth column) shows the successful anticipation of the upcoming peak in the single SEIR wave set of simulations (Fig 5 top row, most of the distribution is situated days before the transition). A similar trend is seen in the set of simulations that had a gradual decrease in *β* (third row), with almost all of the distribution mass occurring over five days before the epidemic transition. In contrast, simulations with an increasing effective contact rate (second row) have the majority of the distribution mass occurring after the re-emergence critical transition, possibly showing a characteristic bifurcation delay seen in a previous COVID-19 analysis [16]. It is unclear whether this is a delayed detection of the critical transition occurring (*R*_0_ = 1) or an early, successful anticipation of the upcoming epidemic transition (*R*_*t*_ = 1) at the peak. Finally, simulations with a step-decrease in *β* (fourth row) behave as expected with the 2-*σ* method showing a delayed detection of the instantaneous forcing through the critical transition. There is no gradual approach to the transition (*β* decreases instantaneously), so critical slowing down should not occur. Similar trends were also seen in the wild-type-like pathogen results, however the lower and later infection curves possibly led to the observed later and more spread-out time-of-detection distributions (S3 Fig).

In the Supporting Information S4 Fig, we also provide a Q–Q plot for the mean-detrended residuals of the simulations at day 10, 50 and 100 for each set of Alpha-like scenarios. We note that for low prevalence values, the residuals show a skewed distribution, with residuals being more normally distributed when prevalence is high. As stated in O’Regan and Drake [12], this is expected because the linear noise approximation assumes a large population limit (which likely does not hold for the *E, I, R* compartments around the disease-free equilibria), and because elimination is an absorbing state. Nonetheless, the simulation variance shows good concordance with the theory under the linear noise approximation, so this approximation is likely useful in deriving the expected trends in second-order statistics of the prevalence time series.

We next plot the corresponding results after applying a negative binomial reporting error with an 80% reporting probability and overdispersion of one Fig 6. The biggest difference in results is that the variance shows monotonic increases up to the epidemic transition. This difference is expected from previous results in the literature, which found analytic differences in the variance in incidence and prevalence for less complex disease models approaching a bifurcation [32, 64]. The time-of-detection distributions are near-identical to those calculated using the prevalence, which indicates the detection method is robust to reporting errors and using incidence data.

**Fig 6.**
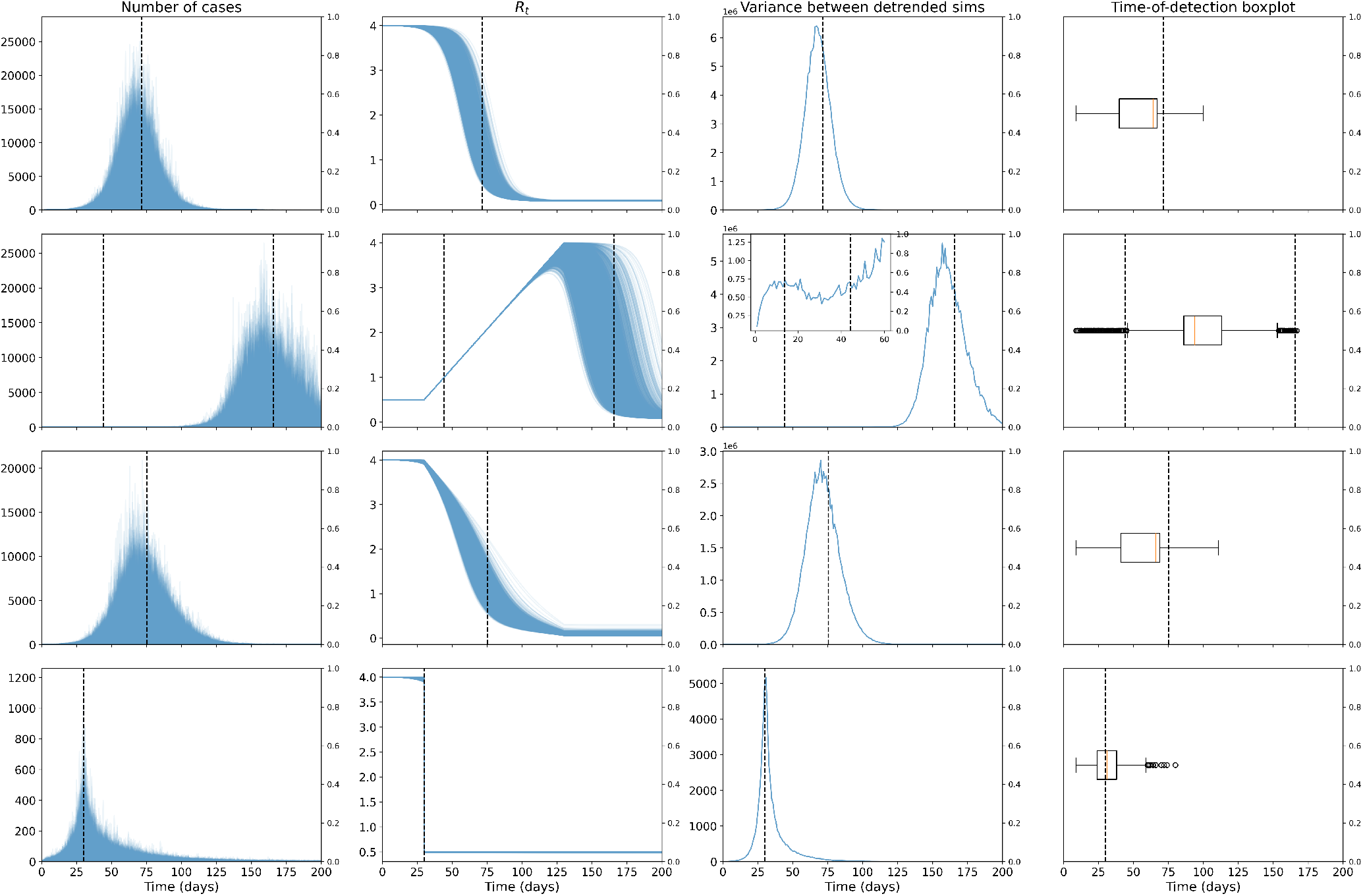
EWS results for the incidence of the Alpha-like pathogen, under a negative binomial reporting error, show successful anticipation of the epidemic transition. Columns show new cases over time, effective reproduction number, variance between the mean-detrended simulations and time-of-detection distribution for the 10,000 simulations run for each of the four modelling scenarios (rows, constant *β*(*t*), increasing *β*(*t*), decreasing *β*(*t*) and a step-decrease in *β*(*t*)) for the Alpha-like pathogen.

The Supporting Information S5 Fig contains similar plots for the other reporting probability and dispersion parameters tested. The shape of the variance curve and time-of-detection distributions are also unchanged under the different parameters.

We note that we assumed no delay in case reporting. Although this is not a realistic assumption, applying a constant reporting delay would only serve to shift all the results by the reporting delay. Thus, the trends in the results would still hold.

### LTLA regional early warning signals

In this section we outline the results using temporal EWS at the LTLA level to predict epidemic transitions in the COVID-19 cases time series between June 2020 and December 2021. We conducted a sensitivity analysis of the rolling window size used to calculate the normalised statistics and also compared the qualitative performance of the normalised signals in providing information of upcoming transitions (see S7 Fig), focusing on the detrended incidence residuals of the Maidstone, Camden and York LTLAs. We also analysed the spatio-temporal trends in early warning signals on the approach to the peak in cases seen in many areas at the end of 2020.

We found that the skew, kurtosis and autocorrelation are noisy for all the LTLAs. Conversely, the index of dispersion and coefficient of variation have too few spikes to provide much predictive power. These observations agree with previous findings that the variance (and related statistics) generally perform best in anticipating transitions [9]. Although using a 10-, 14-, or 30-day window made the resultant time series less noisy, it also smoothed out some of the possible spikes corresponding to a transition detection S7 Fig. We therefore took a rolling window size of seven days and focused on the variance and first difference in the variance in the rest of our analysis of temporal statistics at the LTLA-level case data.

Using the previously outlined method, in the Materials and methods section, we estimated the *R*_*t*_ time series of all LTLAs, and tested the performance of the 2-*σ* method over the COVID-19 pandemic time period considered in terms of anticipating upcoming transitions (Fig 7). There are clearly many 2-*σ* exceedances leading up to the Christmas 2020 peak in the three selected LTLAs as well as the peak in July 2021. There are very few exceedances around April 2021, possibly delayed detections of the epidemic transition around March 2021 in all the LTLAs. The variance appears to have more consistently positive exceedances than the first difference. This is expected as CSD theory predicts increasing variance (and thus more positive exceedances), which would then lead to both positive and negative exceedances in the first difference in variance. The near-zero number of cases in the middle of 2021 also seems to reduce the efficacy of the method (there are far fewer exceedances).

**Fig 7.**
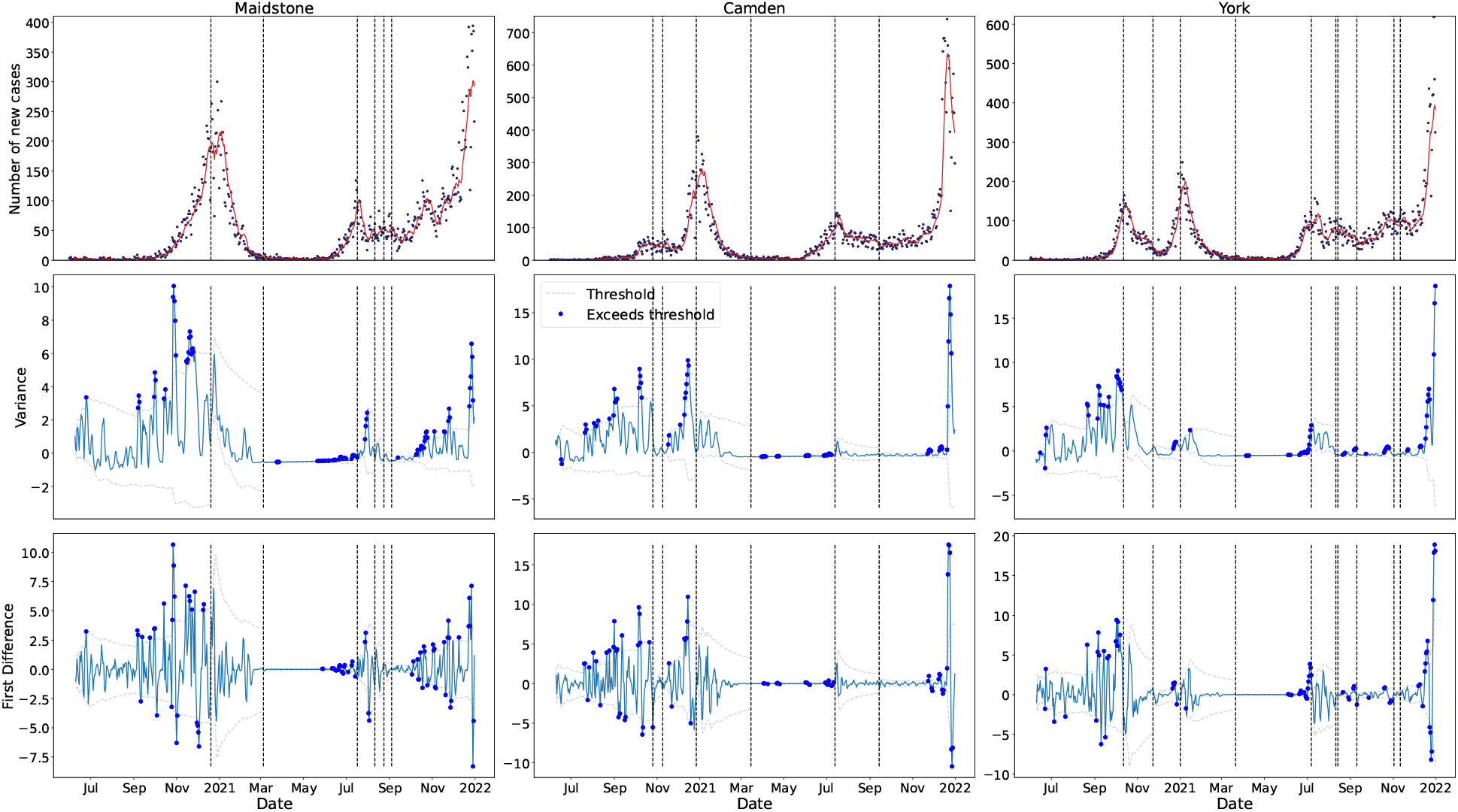
Time series of reported PCR-only COVID-19 cases and selected, normalised time series statistics for a sample of LTLAs between June 2020 and December 2021. The dotted lines show the 2-*σ* threshold, with dots corresponding to time points where the threshold is exceeded. The threshold is recalculated after the approximate times that *R*_*t*_ = 1, shown as the dotted vertical lines. Statistics are calculated using a rolling seven-day window. There are multiple exceedances before the transition point for both signals across all locations.

Next, following the sensitivity analysis in Southall *et al*. [58], we defined the detection time as the time point following at least three consecutive exceedances of the threshold, and calculated all of these occurrences between June 2020 and December 2021 for the three LTLAs of interest (Fig 8).

**Fig 8.**
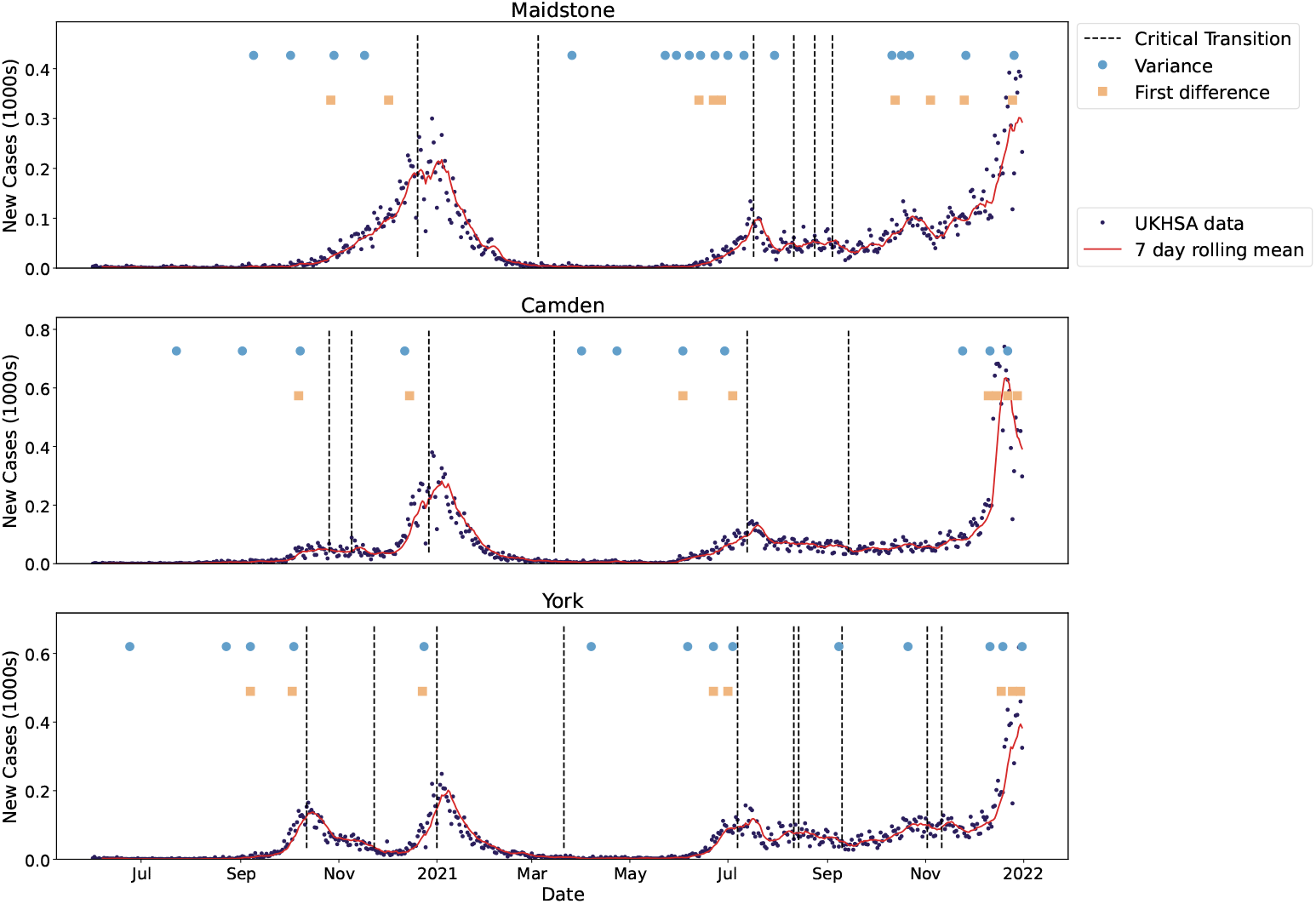
Illustration of the expected epidemic transitions for a sample of LTLAs between June 2020 and October 2021 alongside the times-of-detection for three consecutive threshold exceedances for the top performing EWS (vertical positioning of the signals does not matter). The vertical dotted lines show the expected dates when *R*_*t*_ = 1 and the circles and squares show the time-of-detection for variance and first difference in variance EWS. The incidence of new cases and the seven-day rolling average are also overlaid. The 2-*σ* method successfully anticipates most of the emergence transitions.

Using the variance leads to more robust predictions of the upcoming peaks compared to the first difference in variance, especially for Maidstone, which had a much sharper peak than the other two LTLAs considered. This could be due to the more distinctly positive and consecutive trend in the variance.

The Supporting Information S8 Fig contains sensitivity analysis of these results to changes in rolling window size (for 10, 14 and 30 day windows). As expected, longer window sizes led to fewer and delayed detections. Nonetheless, the results were robust to changes in this window size, with successful detection occurring for all window sizes across all three LTLAs.

Finally, we considered how well the variance anticipates the Christmas 2020 peak in all 315 LTLAs in England. For each LTLA, we considered the times-of-detection that occurred before the Christmas peaks. We discounted any that occurred before the previous critical transition, which mainly corresponded to that wave’s take-off point, or a chosen cut-off point of the end of October as detection before then is likely signalling the take-off, not the peak—e.g. in Camden and York. We now analyse our novel spatio-temporal results for how early each LTLA detected the upcoming peak (Fig 9a), and how soon after the first detection in an LTLA did the other LTLAs signal (Fig 9b).

**Fig 9.**
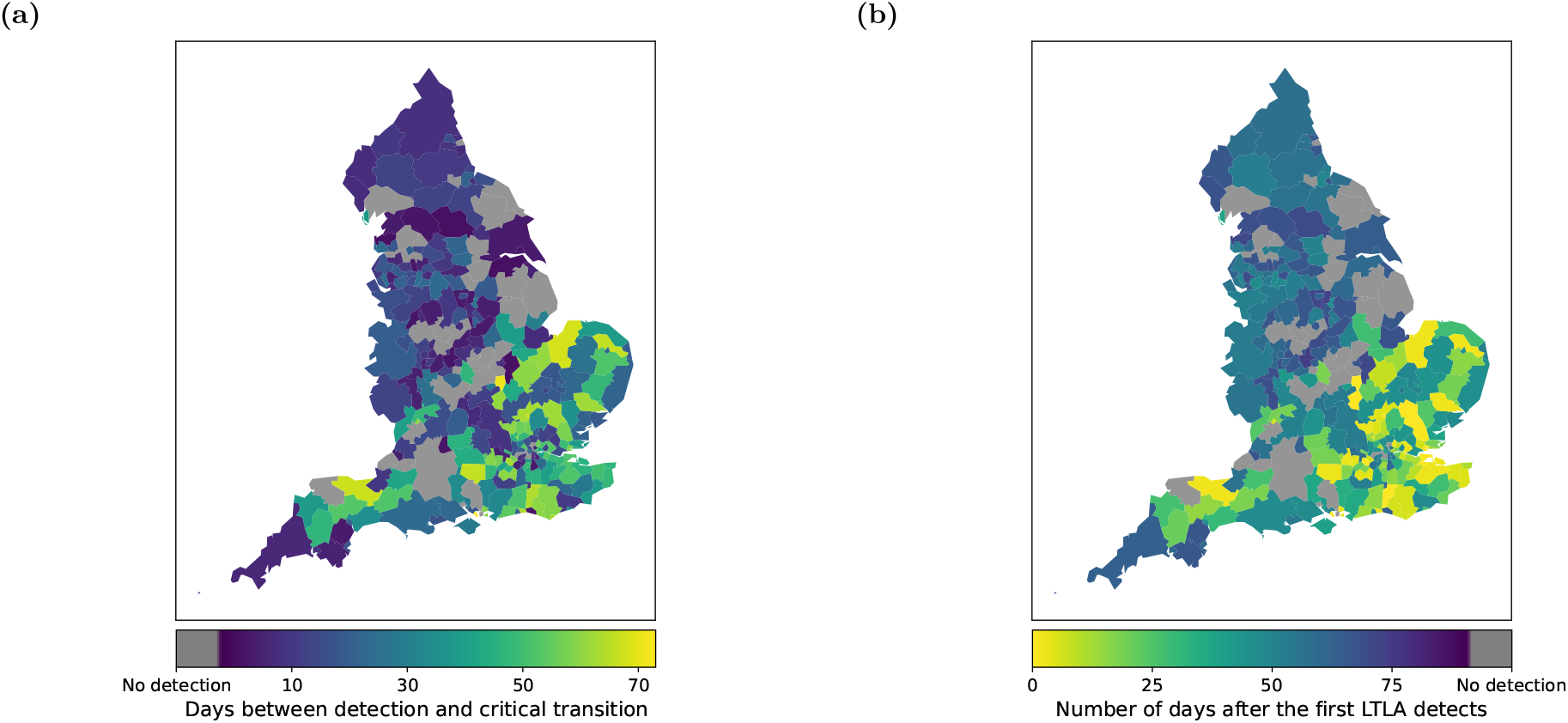
Detection date distributions (relative to the Christmas 2020 peaks) for all 315 LTLAs in England. **(a)** Number of days before the Christmas 2020 peak (for each LTLA) that the upcoming peak is detected (using the variance). **(b)** Number of days after the first day of detection that the upcoming peak is detected for each LTLA. LTLA maps are taken from the Office for National Statistics, available at https://geoportal.statistics.gov.uk/datasets/ons::local-authority-districts-december-2020-boundaries-uk-buc/about. There is a clear spatial trend in the results from the South East to North West of England.

The East and South East of England tended to anticipate the upcoming peak much earlier than the rest of England. With the Alpha-variant thought to have originated in Kent/South East of England, this pattern in signalling could be due to the spread of the Alpha-variant from the South East to the North West of England. Of the 44 LTLAs that did not signal before the peak, most saw ‘flat’ peaks in their reported case time series (or many fluctuations around *R*_*t*_ = 1 at Christmas). The last detecting LTLA (North Warwickshire) occurred 74 days after the first LTLAs that signalled (Milton Keynes, Uttlesford, Gosport, Wellingborough, Runnymede, Mid Sussex and Stevenage).

### NHS regional early warning signals

Having looked at the ability of EWS to predict the critical transitions at the LTLA level, we next investigated their usage for NHS regions. In this analysis, we tested the ability of the spatial variance and spatial skew across the LTLAs in each NHS region to anticipate upcoming peaks and wanes.

Before conducting a corresponding spatio-temporal analysis of NHS regional statistics on the approach to the Christmas 2020 period peaks, we applied the 2-*σ* method to the spatial variance and skew (across each NHS region). The variance clearly exceeds the 2-*σ* threshold more reliably than the skew on approach to critical transitions, full results in S9 Fig. Interestingly, the spatial variance also seems to anticipate, or only slightly lag, the re-emergence wave in mid-2021 (which the temporal EWS at the LTLA level did not reliably identify).

In another set of novel analyses, we tested the ability of the temporal variance in both the new hospitalisations and hospital occupancy time series to anticipate the transitions in the NHS regional cases—to our knowledge, no analysis using hospitalisation data exists in the EWS literature. We applied the 2-*σ* method on the normalised variance to analyse trends in the threshold exceedances (Fig 10, full results in S9 Fig).

**Fig 10.**
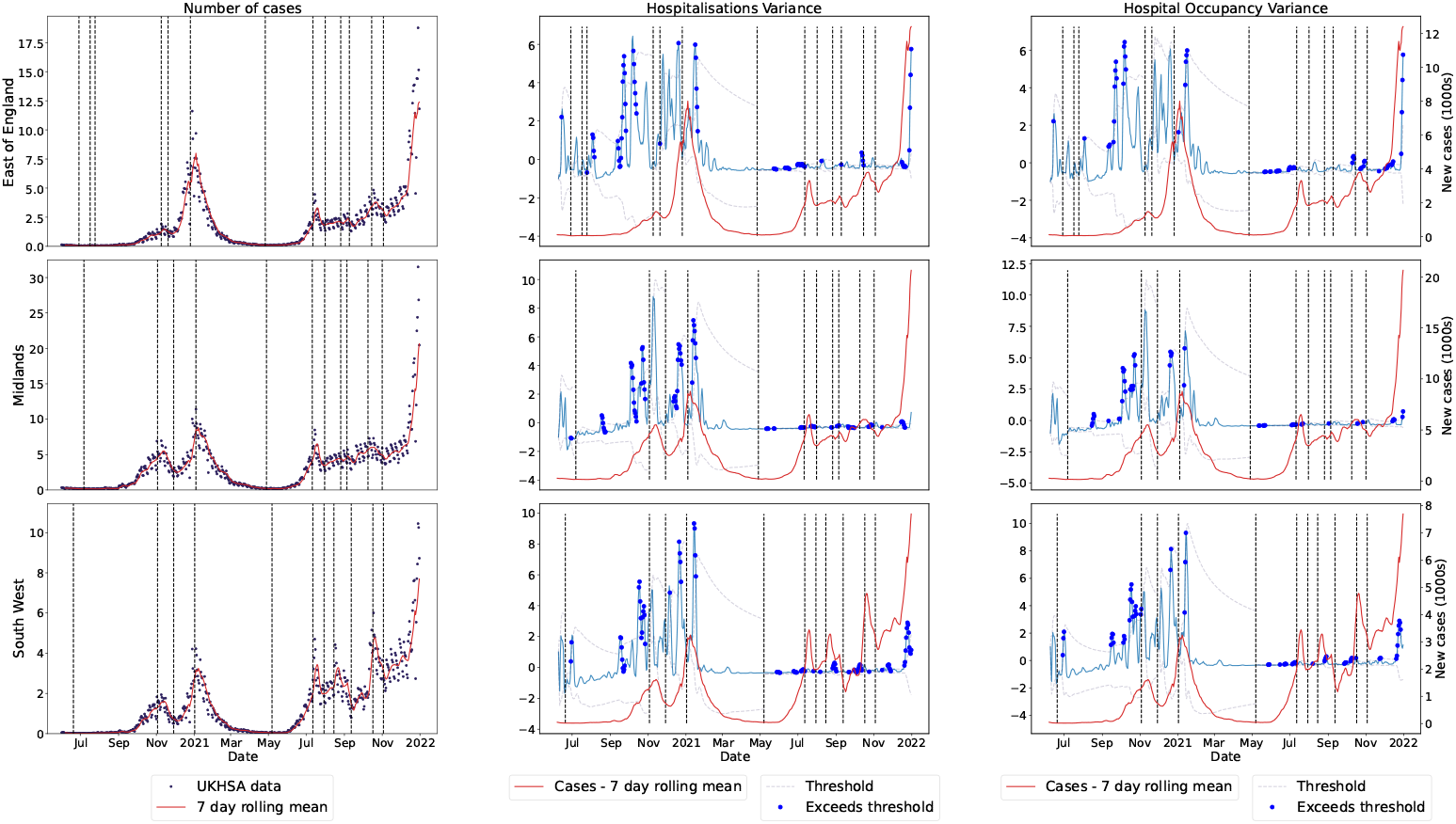
Reported PCR-only COVID-19 cases and selected, normalised temporal statistics calculated over the spatially detrended hospitalisation incidence and hospital occupancy data for the three selected NHS regions between June 2020 and December 2021. The dotted lines show the 2-*σ* threshold, with dots corresponding to time points where the threshold is exceeded. Incidence data is shown as the dark dots, with the red line being the rolling seven-day average. There are clear exceedances before the epidemic transitions in all three NHS regions.

Although hospitalisations and related time series lag infections [65], there are many exceedances upon approach to the epidemic transitions in the NHS-regional level reported case time series. This suggests hospitalisation-based EWS still have predictive power for transitions in the corresponding reported cases time series and could be useful in future epidemic applications.

We conducted a similar timeline analysis into how each of the selected EWS perform in anticipating each of the epidemic transitions identified in the reported case time series for the seven NHS regions (Fig 11).

**Fig 11.**
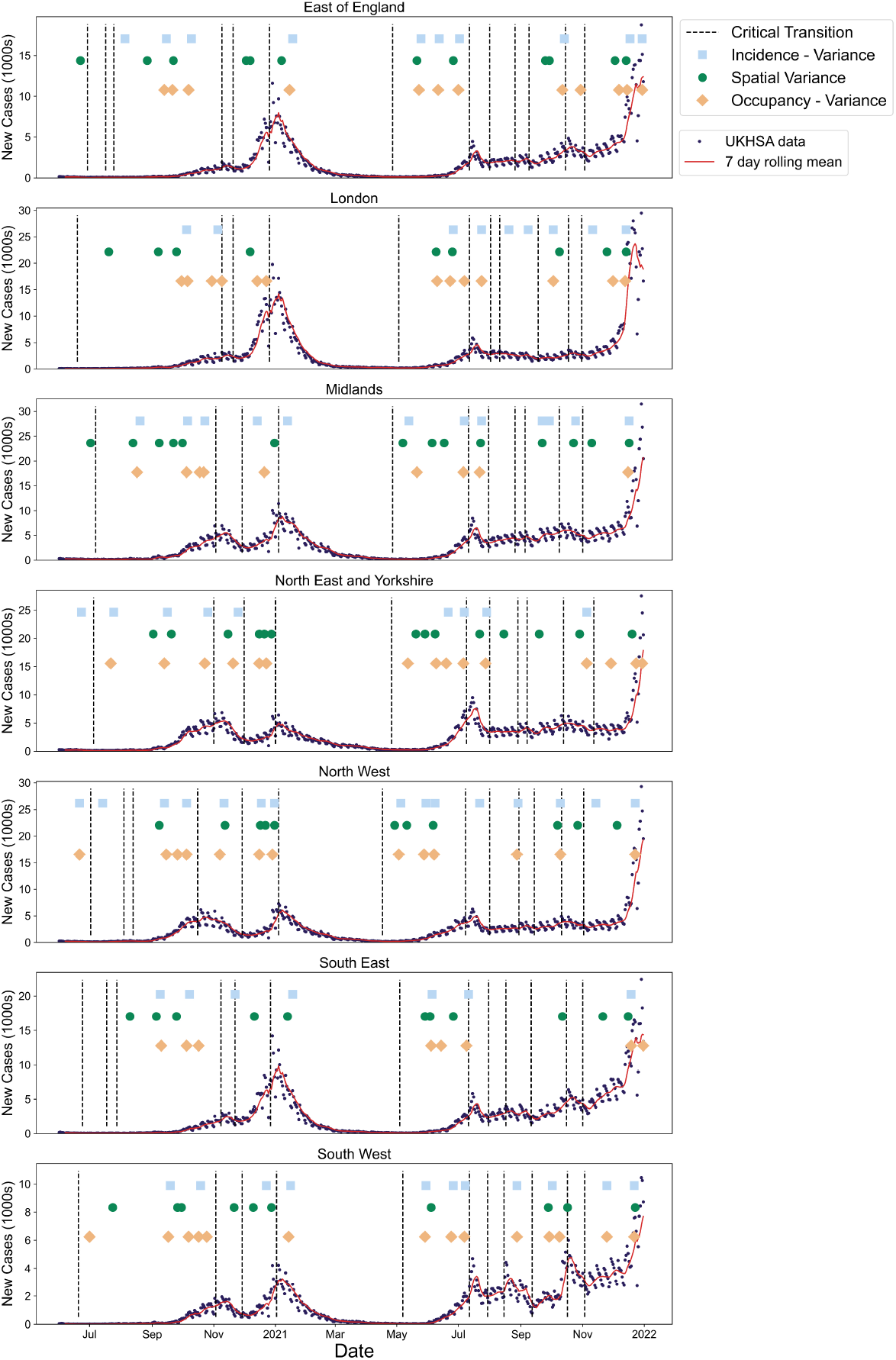
Illustration of the expected epidemic transitions for each of the seven NHS regions between June 2020 and December 2021 alongside the times-of-detection for three consecutive threshold exceedances. The vertical dotted lines show the expected dates when *R*_*t*_ = 1 and the circles, squares and diamonds indicate the time-of-detection for the spatial variance, and variance in hospitalisation-based data. The incidence of new cases and seven-day rolling average are also overlayed. All statistics signal successfully before re-emergence with the spatial variance seemingly the most robust.

Although the spatial variance signals less than the temporal variance in both the new hospitalisations and hospital occupancy, it seems to more reliably anticipate the upcoming peaks. Considering the peaks near the start of November 2020, January 2021 and July 2021 for all NHS regions, the sensitivity of the spatial variance was highest (at one), followed by the variance in incidence (0.857) and then variance in hospital occupancy (0.810). The spatial variance also seems to signal earlier than the temporal metrics, although the efficacy of each differs by region. Nonetheless, all EWS show multiple detections upon approach to almost all of the peaks and troughs in each region.

Finally, we considered how well the spatial variance anticipates the Christmas 2020 peak in each NHS region using the same method as the temporal variance with the LTLAs (Fig 12).

**Fig 12.**
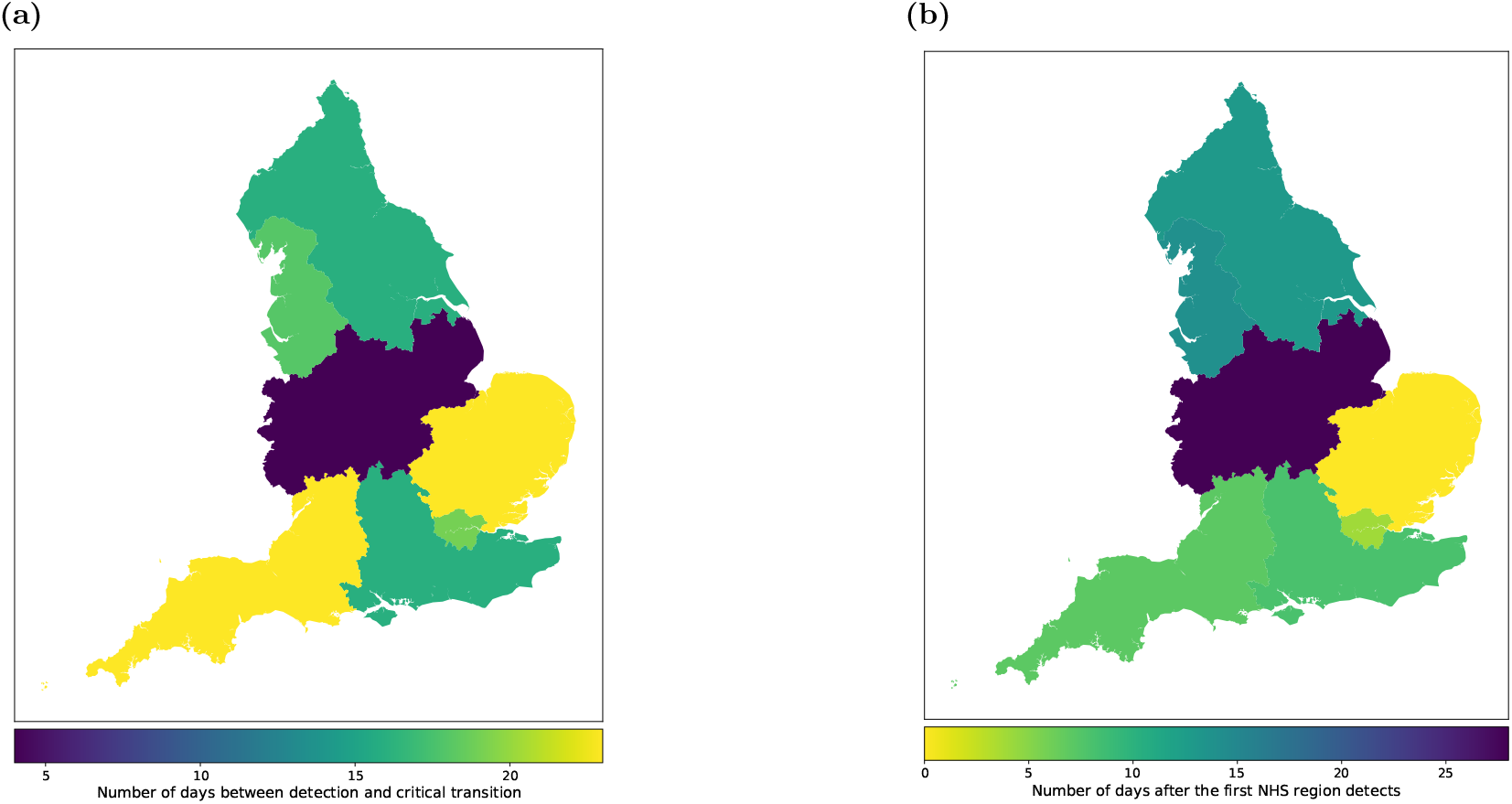
Detection date distributions (relative to the Christmas 2020 peaks) for all seven NHS regions in England. **(a)** Number of days before the Christmas 2020 peak (for each NHS region) that the upcoming peak is detected (using the variance). **(b)** Number of days after the first day of detection that the upcoming peak is detected for each NHS region. NHS maps taken from the Office for National Statistics, available at https://geoportal.statistics.gov.uk/datasets/ons::nhs-england-regions-april-2020-boundaries-en-buc/about. Similar trends are seen in the spatial pattern as with the LTLA results.

The East of England is the earliest NHS region to signal with a time-of-detection 23 days before its Christmas epidemic transition. This mirrors the pattern seen in the LTLA results. Although the Midlands signals last, it still anticipates the peak by four days. Regions where the corresponding LTLAs had a very early time-of-detection (Fig 9b) are also quick to signal. However, the South West, which had some poor predictions at the LTLA level, also signals first.

## Discussion

This paper analyses the ability of early warning signals to predict epidemic transitions and non-stationary periods of the reported cases time series in England for the June 2020 to December 2021 period of the COVID-19 pandemic. Our analysis considered both temporal and spatial statistics of LTLA and NHS regional case data. In addition, we extended the literature on EWS under SEIR-like contagion dynamics through a theoretical and simulation-based analysis of both prevalence and incidence data. Finally, we also investigated whether EWS analysis of hospitalisation data can identify transitions in corresponding disease time series.

We built on existing literature showing the ability of EWS to anticipate transient dynamics in disease time series for diseases with a latent period and case reporting errors by using *β*(*t*) as the driving parameter and considering a possible analytic framework for the transient dynamics (Figs 5 and 6). At the time of writing, this mirrors the only previous EWS analyses of the SEIR model in the literature (Brett *et al*. [25] and Dablander *et al*. [16]). Brett et al. [25] established that SEIR-based disease models of increasing complexity show signs of CSD and Dablander *et al*. [16] considered how EWS of re-emergence may be subject to bifurcation delays, depending on how *β*(*t*) was increased [66]. Nonetheless, our results are novel in using *β*(*t*) as the driving parameter to model different scenarios likely seen in the course of the COVID-19 pandemic. To the best of the authors’ knowledge, we are also the first to compare the analytical time-dependent variance to numerical results for the SEIR model. The observed two peaks in the variance shows the importance of considering transient disease dynamics and agrees with previous results that CSD can be observed before non-linear periods (not just critical transitions) and is affected by overlapping disease time scales (in this case a change in the linearised dynamics near the disease-free equilibrium compared to the rest of the disease trajectory) [16, 22, 23]. Using this, we were also the first to show that the *R*_*t*_ = 1 transition at the peak can be successfully anticipated. Future study could further develop the theory of transient indicators along a disease trajectory (such as recent developments in [63]).

Conversely, the increasing *β* results indicate potential problems in applying EWS to disease dynamics with multiple transitions and time scales (as previously considered in [16]). Only 15% of the (re-)emergence simulations signalled before the critical transition, and with prevalence so low at the start of the simulations, these are unlikely to be caused by dynamic instability and instead due to the low proportion of infections [16]. Most of the time-of-detection mass occurred between the two transitions (Figs 5 and 6 fourth columns), which decreases the interpretability of the detections. These may be successful anticipations or delayed detections; the EWS alone provide no information about the time or type of the upcoming transition. Finally, Brett *et al*. [25] used a different driving parameter (a decrease in the vaccination rate) and successfully anticipated the re-emergence transition. Future work should investigate whether this discrepancy is due to the difference in the model or in the disease time scales.

Our results also show the robustness of detection to case reporting errors. Mirroring the prevalence results, the time-of-detection distributions for incidence results pre-empted the epidemic transitions. Further, these results also highlight the differences in EWS trends between prevalence- and incidence-based statistics as previously investigated in [32, 64]. Specifically, the double peak in variance, due to the two changes in complexity of the time-dependent Jacobian eigenvalues, is not present in the incidence variance; the incidence variance shows a monotonic increase upon approach to the dynamic transition point. As mentioned in [18], future work could investigate the analytic results for different case reporting error distributions. Although our results were robust to reporting errors, we note that the assumption of a constant reporting probability, dispersion and reporting delay are unlikely to hold in a real disease outbreak. Future work could also investigate the effects of the reporting error distribution parameters being time-dependent and correlated with the prevalence level.

The COVID-19 results indicate that EWS analysis could have reduced the uncertainty in the timing and effectiveness of intervention methods around the Christmas 2020 peak (Fig 9) [67]. The successful detection of the Christmas peak in over 86% of the LTLAs indicates that our proposed EWS analysis might have helped guide policy decisions on the duration and location of NPIs. Further, using the 2-*σ* method may have provided early warning of the introductions of the Alpha and Delta waves, as they signalled well in advance of the re-infection waves. However, at both LTLA level and NHS regional level seemingly no region anticipated the *R*_*t*_ = 1 transition near the return to endemicity in March 2021. This again highlights the problem with applying EWS in practice; although the theory of CSD and EWS is clear, interpreting the signals in an online manner is difficult without further context or modelling. Nevertheless, most regions successfully indicated the non-stationary/increasing periods in reported cases in October-December 2020 and July 2021 which still shows the benefits of including EWS alongside other modelling efforts.

Our spatial analysis reinforces prior results in the literature that spatio-temporal trends should be considered in early warning signals [27, 54, 55, 57]. The spatial variance performed well as a signal at the NHS regional level, anticipating the majority of the waves in the reported case time series. The spatio-temporal maps (Figs 9 and 12) also illustrate that considering the spread of detections across a country or region may provide extra information to modellers. The early signalling of the East and South East of England may have helped modellers identify the emergence of the Alpha variant and its subsequent spatial spread.

Our novel analysis of hospitalisation-based signals indicates their potential usefulness for future epidemics (Fig 11). They performed similarly to the spatial statistics in prediction power, but also have their own inherent limitations. Hospitals may face social or operating pressures to reduce occupancy rates, potentially causing bias. Further, hospitalisation-based statistics may be skewed by operating capacity in hospitals. Such external factors can limit EWS efficacy. Future work could consider using previously proposed composite metrics to investigate whether combining statistics of both incidence and hospitalisation time series leads to better results [7, 58].

In this article, we have investigated the application of EWS to anticipate epidemic transitions through both a simulation-based and data-driven analysis. These developments highlight the applicability of including EWS in real-time epidemic modelling alongside more traditional modelling efforts.

## Supporting Information

**S1 Text. SIS Derivations**.

**S2 Fig. Proportion infectious under different modelling scenarios**. Proportion infectious over time for the ten thousand simulations run for each of the four modelling scenarios (constant *β*(*t*), increasing *β*(*t*), decreasing *β*(*t*) and a step-decrease in *β*(*t*)). (a) Alpha-like pathogen. (b) Wild-type-like pathogen.

**S3 Fig. Early warning signal results for a wild-type-like pathogen**. Proportion infectious over time, effective reproduction number, variance between the mean-detrended simulations and time-of-detection distribution for the ten thousand simulations run for each of the four modelling scenarios (constant *β*(*t*), increasing *β*(*t*), decreasing *β*(*t*) and a step-decrease in *β*(*t*)) for the wild-type-like pathogen.

**S4 Fig. Q–Q Plots for the Alpha-variant-like simulations**. Plots for each of the four modelling scenarios (constant *β*(*t*), increasing *β*(*t*), decreasing *β*(*t*) and a step-decrease in *β*(*t*)) at three different time points.

**S5 Fig. Early warning signal results for Alpha-like simulations under different case reporting distributions**. For all figures: reported cases, effective reproduction number, variance between the mean-detrended simulations and time-of-detection distribution for the ten thousand simulations run for each of the four modelling scenarios (constant *β*(*t*), increasing *β*(*t*), decreasing *β*(*t*) and a step-decrease in *β*(*t*)). **S5A: 80% reporting probability and dispersion of 10. S5B: 60% reporting probability and dispersion of 1. S5C: 60% reporting probability and dispersion of 10**.

**S6 Fig. Timeline of the sequenced variants of COVID-19 in the UK between June 2020 and December 2021**. Distribution shown is the proportion of total sequences represented by each (named) variant.

**S7 Fig. Sensitivity analysis of possible LTLA time series statistics to rolling window size**. All figures plot the time series of new COVID-19 cases and selected time series statistics for a sample of LTLAs between June 2020 and December 2021. **S7A: Normalised statistics with a 7-day and 14-day window. S7B: Statistics with a 10-day and 30-day window**.

**S8 Fig. Sensitivity analysis of the 2-***σ* **method applied to LTLA time series to the size of the rolling window**. All figures show the expected epidemic transitions for a sample of LTLAs between June 2020 and October 2021 alongside the times-of-detection for three consecutive threshold exceedances for the top performing EWS with the incidence of new cases overlayed. **S8A: 10-day window. S8B: 14-day window. S8C: 30-day window**.

**S9 Fig. Normalised NHS time series statistics for all NHS regions**. ALl figures plot the incidence and normalised statistics with expected epidemic transition points and 2-*σ* exceedances. **S9A: Statistics for reported cases. S9B: Statistics for hospitalisation incidence and hospitalised cases**.

## Author contributions

**Joshua Looker:** Data curation, Formal analysis, Investigation, Methodology, Software, Validation, Visualisation, Writing - Original Draft, Writing - Review & Editing. **Kat Rock:** Conceptualisation, Supervision, Visualisation, Writing - Review & Editing. **Louise Dyson:** Conceptualisation, Supervision, Visualisation, Writing - Review & Editing.

## Data availability

All data utilised in this study are publicly available, with relevant references and data repositories provided. The base LTLA maps and NHS maps are available at https://geoportal.statistics.gov.uk/datasets/ons::local-authority-districts-december-2020-boundaries-uk-buc/about, https://geoportal.statistics.gov.uk/datasets/ons::nhs-england-regions-april-2020-boundaries-en-buc/about under Open Government License 3.0.

## Code availability

The code repository for the study is available at: https://github.com/joshlooks/COVIDews

Archived code available at: https://doi.org/10.5281/zenodo.14852091

