## Supplementary material for "Identifying COVID-19 peaks using early warning signals": S1 Text

### SIS Model Equations

The compartment diagram for the SIS model is given in Fig. S1 below. Previous analyses have considered an immigration force of infection ($\eta$), but we instead considered the effective transmission rate ($\beta$) between susceptible ($S$) and infectious ($I$) individuals as the control parameter [1–3].

Assuming a constant population size of $N$ (such that there is no demography/births and deaths), this model, which has $R_0 = \beta/\gamma$ (where $\gamma$ is the recovery rate), has two fixed points: a disease-free equilibrium ($S^* = N$); and an endemic equilibrium ($S^* = N/R_0 = N\gamma/\beta, I^* = N(1 - 1/R_0)$). The stability of each is also given by $R_0$, with the endemic equilibrium (disease-free equilibrium) being stable (unstable) for $R_0 > 1$ and unstable (stable) for $R_0 < 1$.

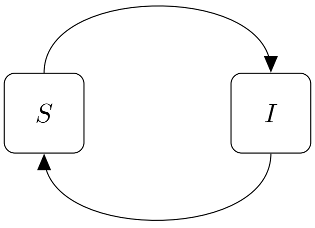


**Fig S1.** SIS compartmental diagram showing the infection and recovery processes. Transmission occurs at a rate of $\beta$, and recovery occurs at a rate of $\gamma$.

The corresponding system is then one-dimensional (assuming a constant population size such that $S + I = N$, where $N$ is the total population size) with the ordinary differential equation (ODE) for $I$ as:

$$\frac{\mathrm{d}I}{\mathrm{d}t} = \beta(N - I)I/N - \gamma I. \tag{S1}$$

We use the system size expansion and linear noise approximation to express the relationship:

$$I = N\phi(t) + N^{1/2}\zeta, \tag{S2}$$

between $I$, its expected proportion $\phi(t) = \langle I \rangle/N$ and the deviations, $\zeta(t)$, from this proportion (as expected from the law of large numbers [4]).

To first order (in $N$), the system can then be expressed as:

$$\frac{\mathrm{d}\phi}{\mathrm{d}t} = \beta\phi(1 - \phi) - \gamma\phi. \tag{S3}$$

The Jacobean of this system is found as: $J = \beta(1 - 2\phi) - \gamma$ and the noise-covariance matrix is given by $B = \beta\phi(1 - \phi) - \gamma\phi$. Thus, we can find the FPE and corresponding SDE as:

$$\frac{\partial \Pi(\zeta, t)}{\partial t} = (\beta(1 - 2\phi) - \gamma)\frac{\partial \zeta \Pi}{\partial \zeta} + \frac{1}{2}(\beta\phi(1 - \phi) - \gamma\phi)\frac{\partial^2 \Pi}{\partial \zeta^2}, \tag{S4}$$

$$d\zeta = (\beta(1 - 2\phi) - \gamma)\zeta dt + \sqrt{\beta\phi(1 - \phi) + \gamma\phi}dW_t, \tag{S5}$$

agreeing with previous results [1, 2, 5].

Taking the moments of the FPE, the variance and other statistics can then be calculated.
