## Supplementary material for "Identifying COVID-19 peaks using early warning signals": S2 Figure

### Simulation Trajectories

(a) (b)

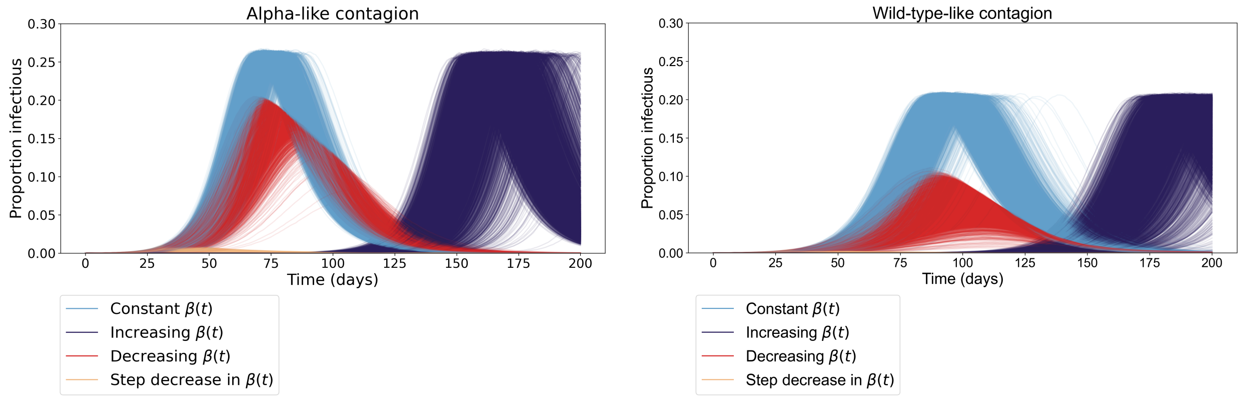


**Fig S2A.** Proportion infectious over time for the ten thousand simulations run for each of the four modelling scenarios (constant $\beta(t)$, increasing $\beta(t)$, decreasing $\beta(t)$ and a step-decrease in $\beta(t)$). **(a)** Alpha-like pathogen. **(b)** Wild-type-like pathogen.
