## Supplementary material for "Identifying COVID-19 peaks using early warning signals": S3 Figure

### Early warning signals for a Wild-type-like pathogen

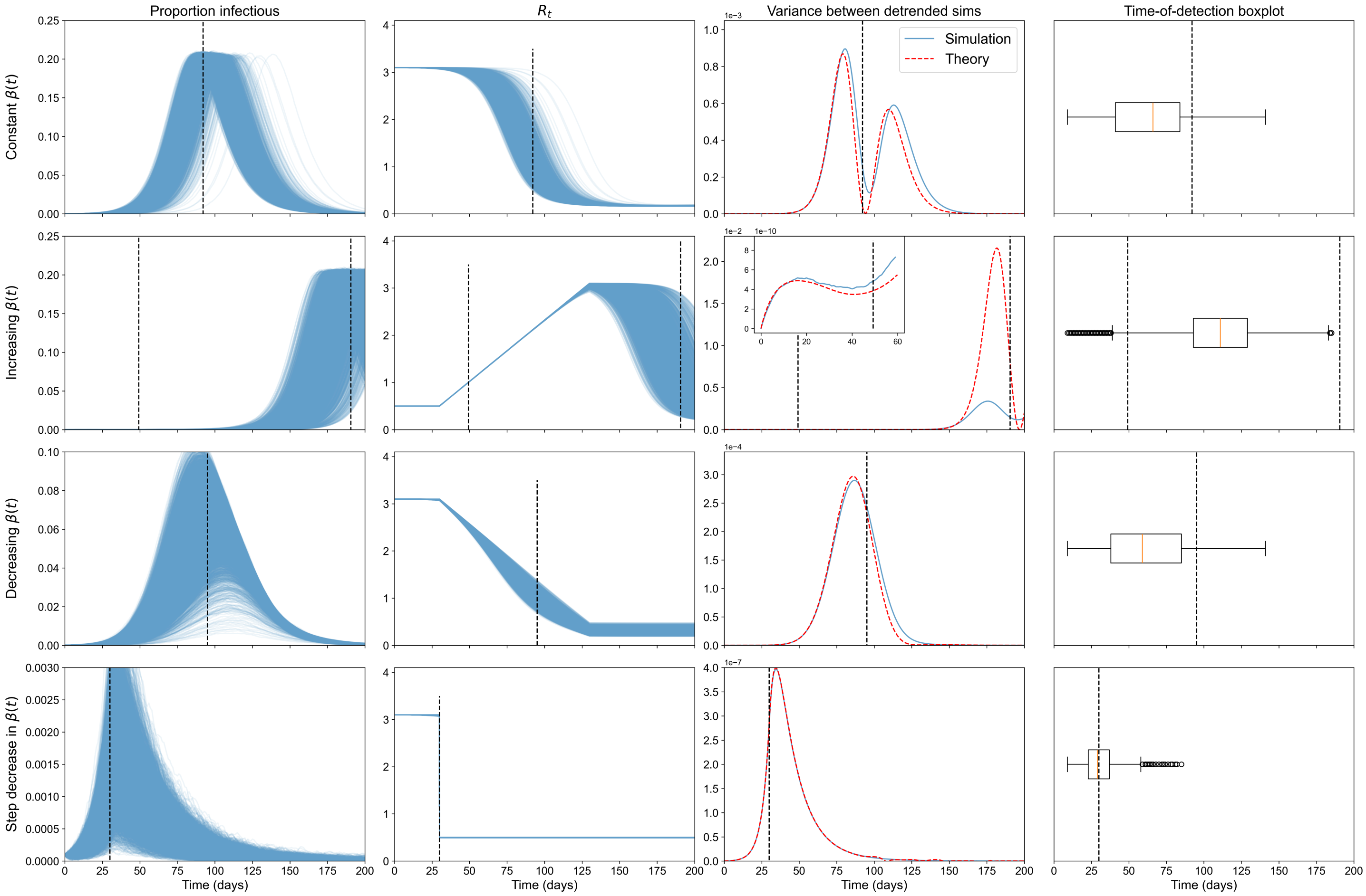


**Fig S3A.** Proportion infectious over time, effective reproduction number, variance between the mean-detrended simulations and time-of-detection distribution for the ten thousand simulations run for each of the four modelling scenarios (constant $\beta(t)$, increasing $\beta(t)$, decreasing $\beta(t)$ and a step-decrease in $\beta(t)$) for the wild-type-like pathogen.
