## Supplementary material for "Identifying COVID-19 peaks using early warning signals": S4 Figure

### Q–Q Plots

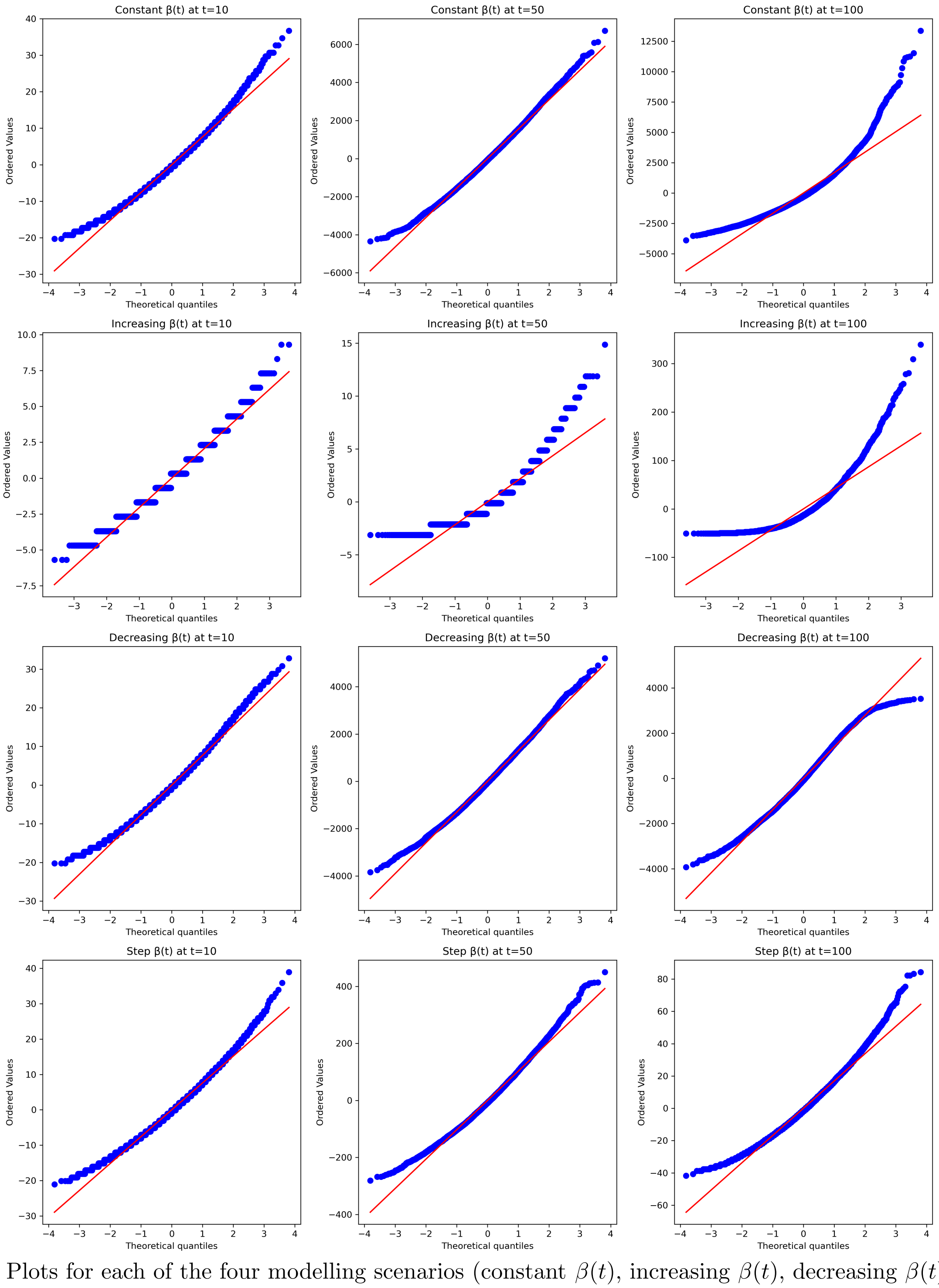


**Fig S4A.** Q–Q Plots for each of the four modelling scenarios (constant $\beta(t)$, increasing $\beta(t)$, decreasing $\beta(t)$ and a step-decrease in $\beta(t)$) at three different time points.
