## Supplementary material for "Identifying COVID-19 peaks using early warning signals": S5 Figure

### Sensitivity analysis for case reporting distribution

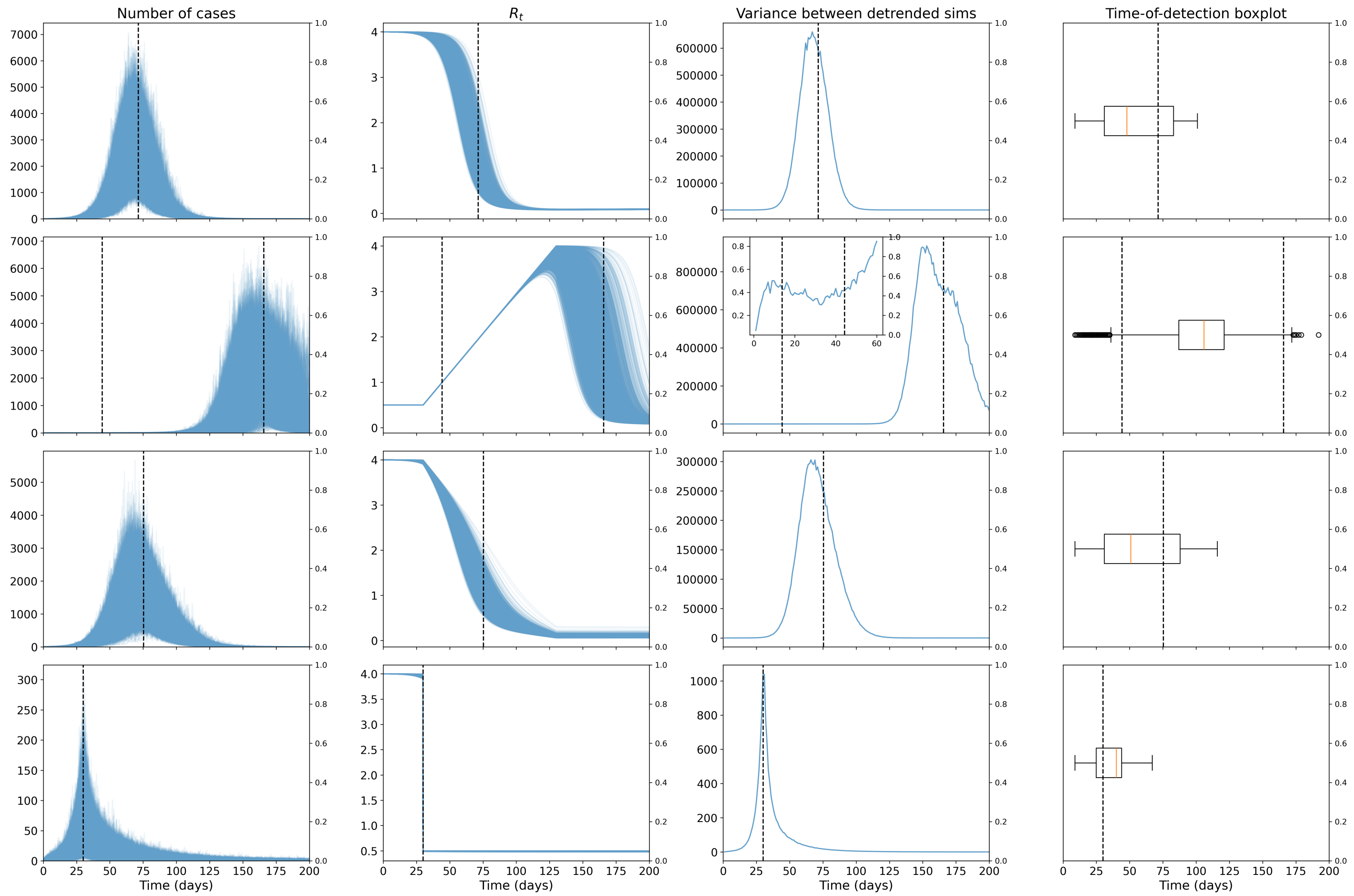


**Fig. S5A.** Reported cases, effective reproduction number, variance between the mean-detrended simulations and time-of-detection distribution for the ten thousand simulations run for each of the four modelling scenarios (constant $\beta(t)$, increasing $\beta(t)$, decreasing $\beta(t)$ and a step-decrease in $\beta(t)$) for an Alpha-like pathogen with an 80% reporting probability and dispersion of 10.

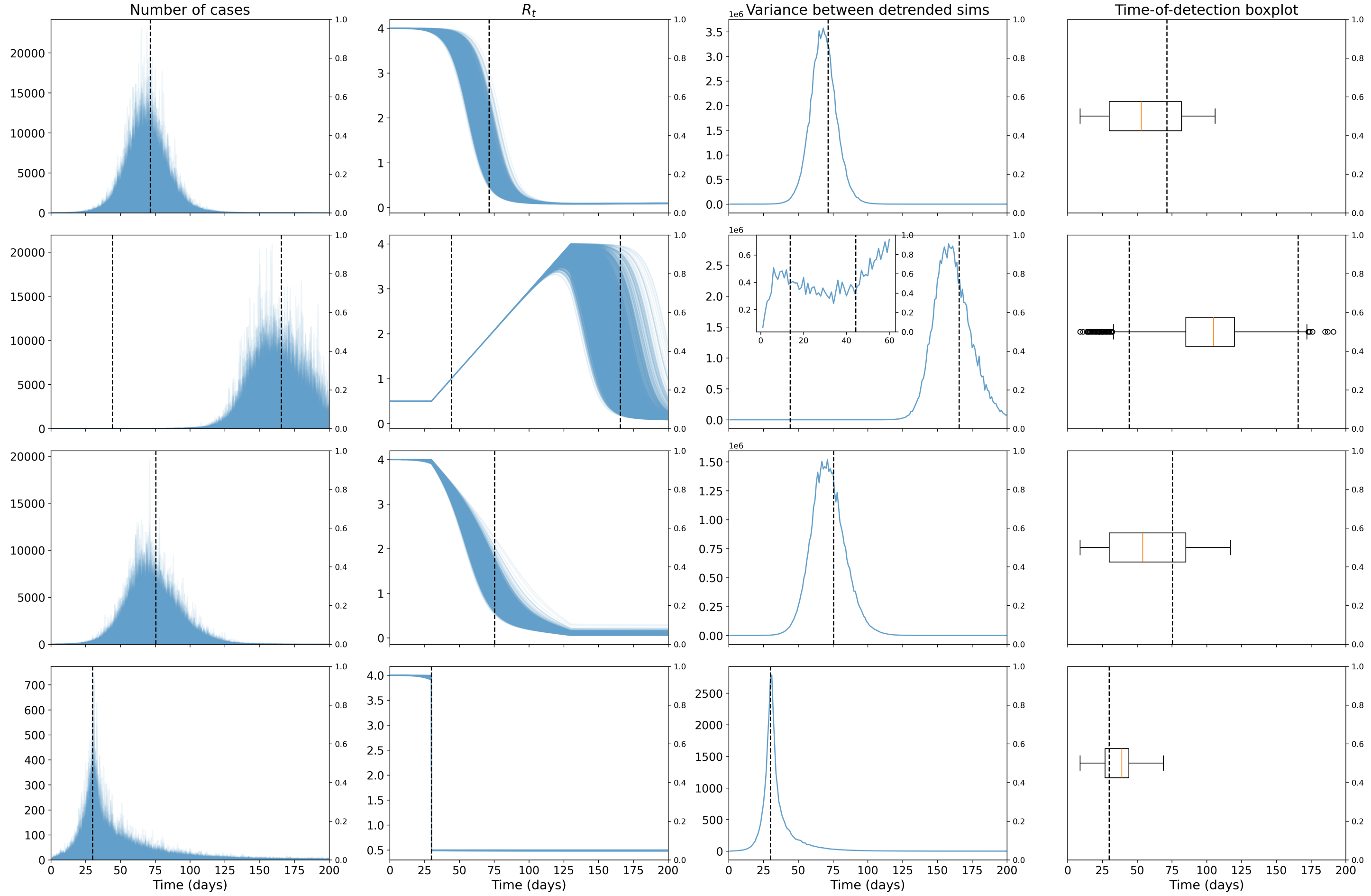


**Fig. S5B.** Reported cases, effective reproduction number, variance between the mean-detrended simulations and time-of-detection distribution for the ten thousand simulations run for each of the four modelling scenarios (constant $\beta(t)$, increasing $\beta(t)$, decreasing $\beta(t)$ and a step-decrease in $\beta(t)$) for an Alpha-like pathogen with a 60% reporting probability and dispersion of 1.

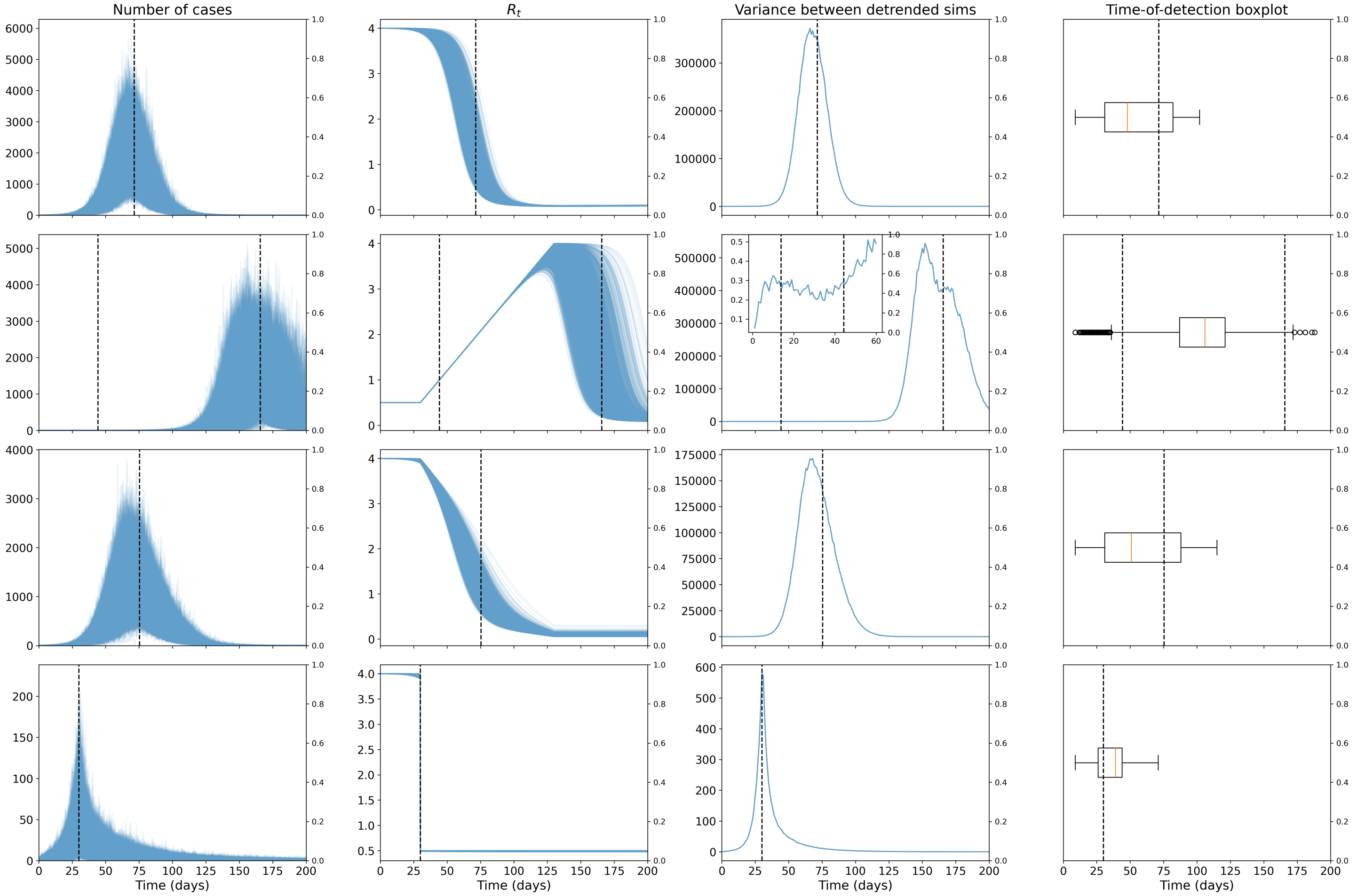


**Fig. S5C.** Reported cases, effective reproduction number, variance between the mean-detrended simulations and time-of-detection distribution for the ten thousand simulations run for each of the four modelling scenarios (constant $\beta(t)$, increasing $\beta(t)$, decreasing $\beta(t)$ and a step-decrease in $\beta(t)$) for an Alpha-like pathogen with a 60% reporting probability and dispersion of 10.
