## Supplementary material for "Identifying COVID-19 peaks using early warning signals": S6 Figure

### Variants Timeline

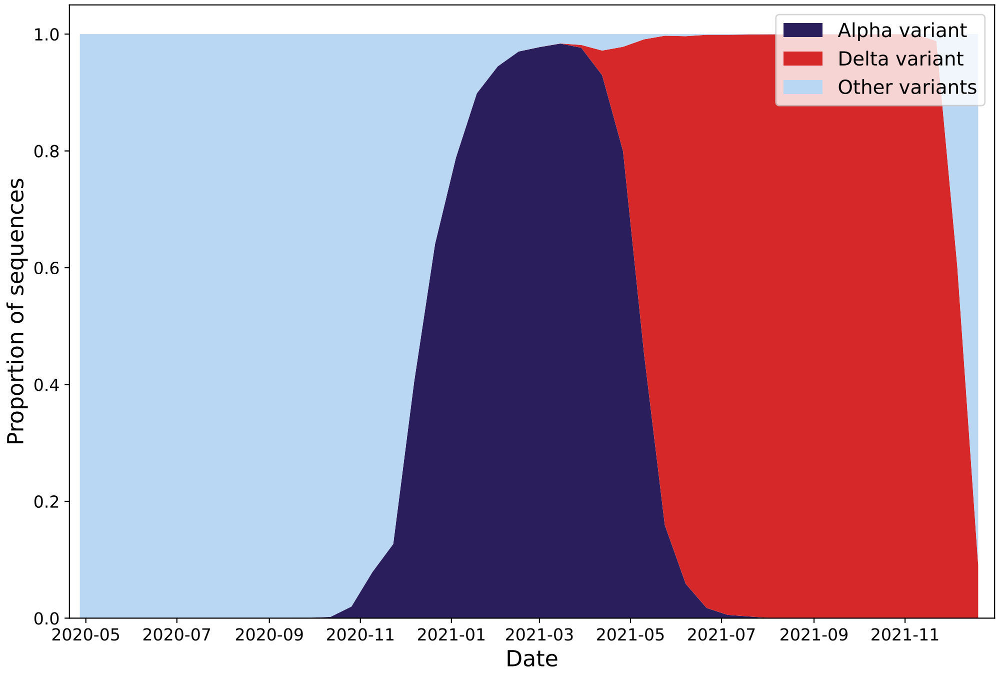


**Fig. S6A.** Timeline of the sequenced variants of COVID-19 in the UK between June 2020 and December 2021. Distribution shown is the proportion of total sequences represented by each (named) variant.
