## Supplementary material for "Identifying COVID-19 peaks using early warning signals": S7 Figure

### Sensitivity analysis of time series statistics to window size

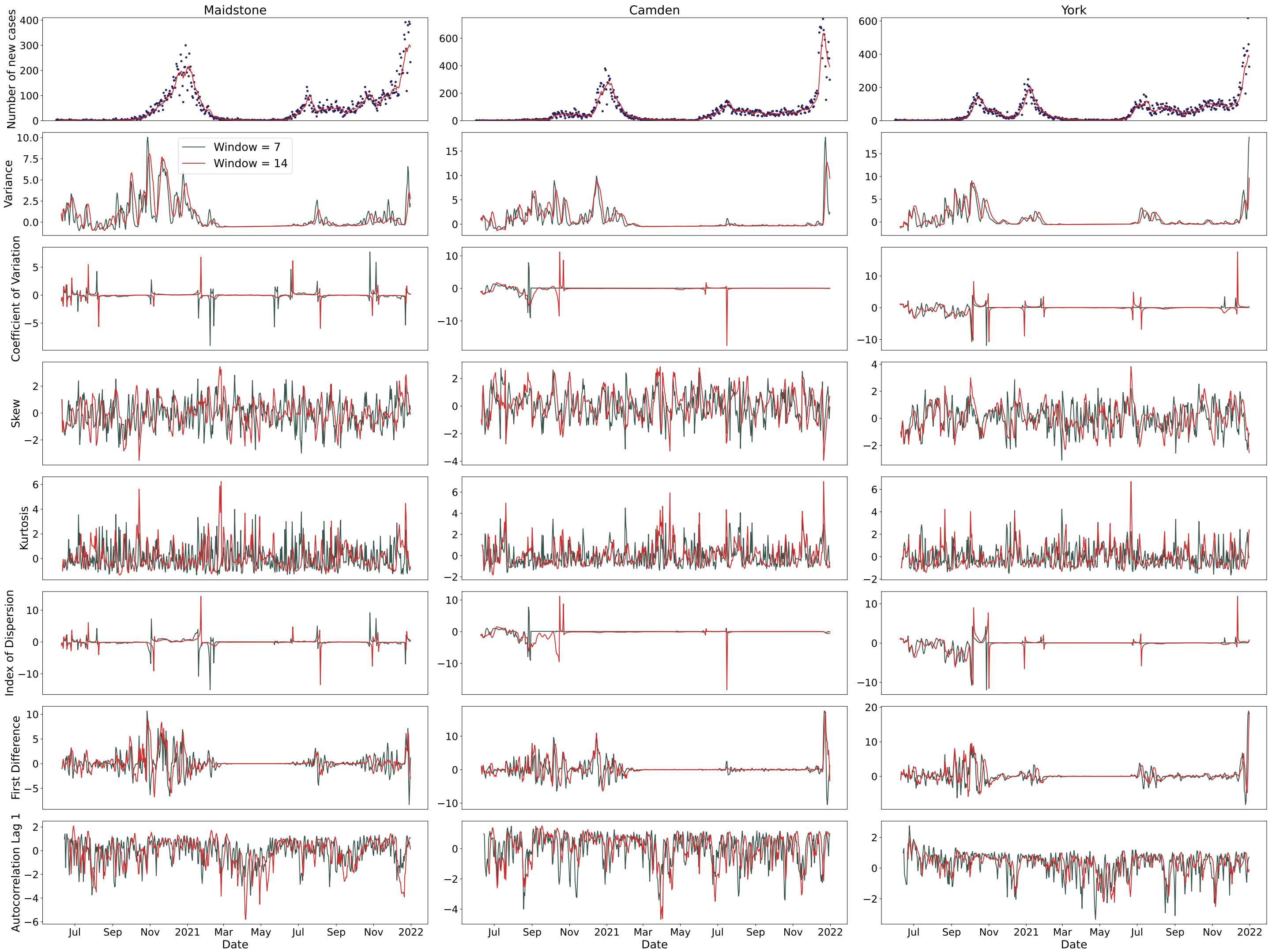


**Fig. S7A.** Time series of new COVID-19 cases and selected normalised time series statistics for a sample of LTLAs between June 2020 and December 2021. Statistics are calculated using a rolling 7- or 14-day window.

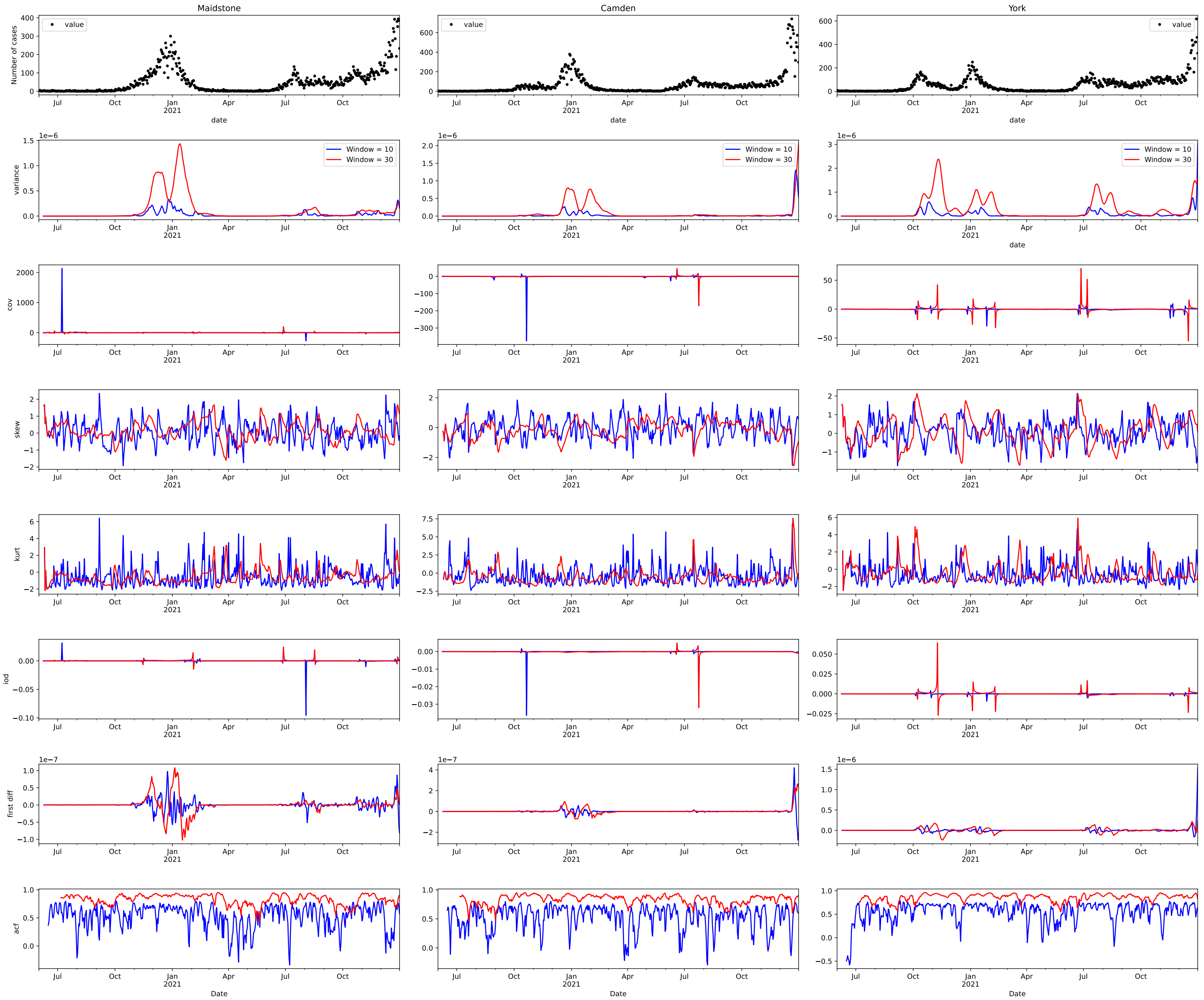


**Fig. S7B.** Time series of new COVID-19 cases and selected time series statistics for a sample of LTLAs between June 2020 and December 2021. Signals were calculated using a rolling 10- or 30-day window.
