## Supplementary material for "Identifying COVID-19 peaks using early warning signals": S8 Figure

### Sensitivity analysis of 2-$\sigma$ to window size for LTLAs

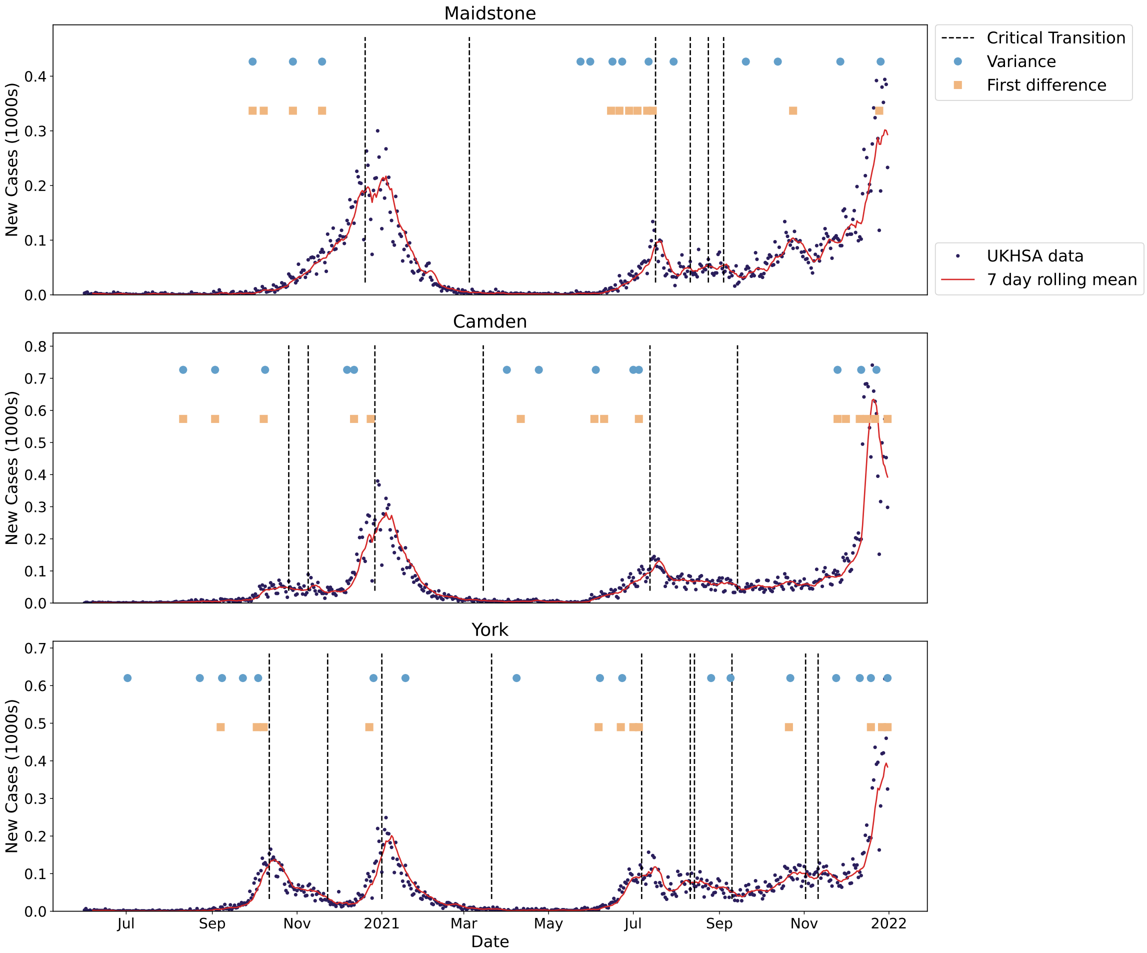


**Fig. S8A.** Illustration of the expected epidemic transitions for a sample of LTLAs between June 2020 and October 2021 alongside the times-of-detection for three consecutive threshold exceedances for the top performing EWS (vertical positioning of the signals does not matter) using a rolling window size of 10. The vertical dotted lines show the expected dates when $R_t = 1$ and the circles and squares show the time-of-detection for variance and first difference in variance EWS. The incidence of new cases and seven-day rolling average are also overlaid.

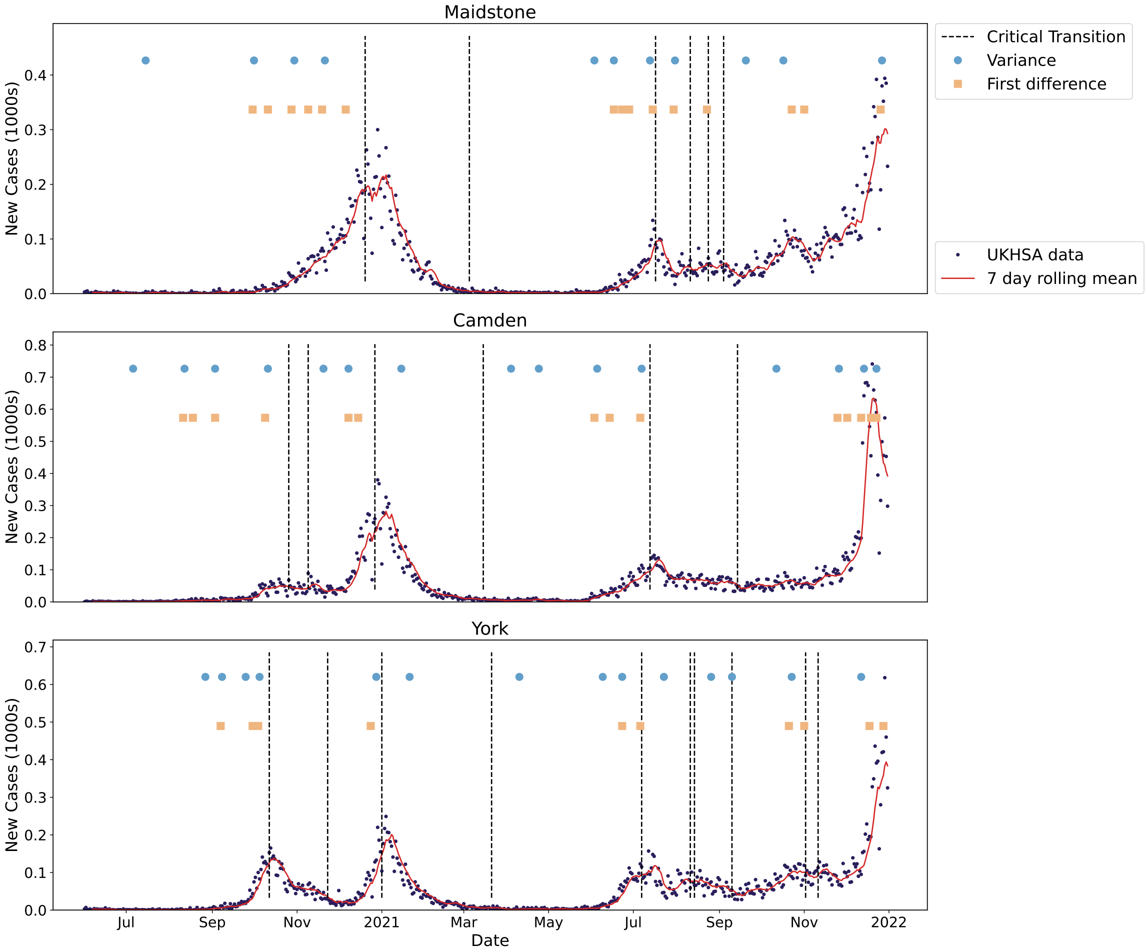


**Fig. S8B.** Illustration of the expected epidemic transitions for a sample of LTLAs between June 2020 and October 2021 alongside the times-of-detection for three consecutive threshold exceedances for the top performing EWS (vertical positioning of the signals does not matter) using a rolling window size of 14. The vertical dotted lines show the expected dates when $R_t = 1$ and the circles and squares show the time-of-detection for variance and first difference in variance EWS. The incidence of new cases and seven-day rolling average are also overlaid.

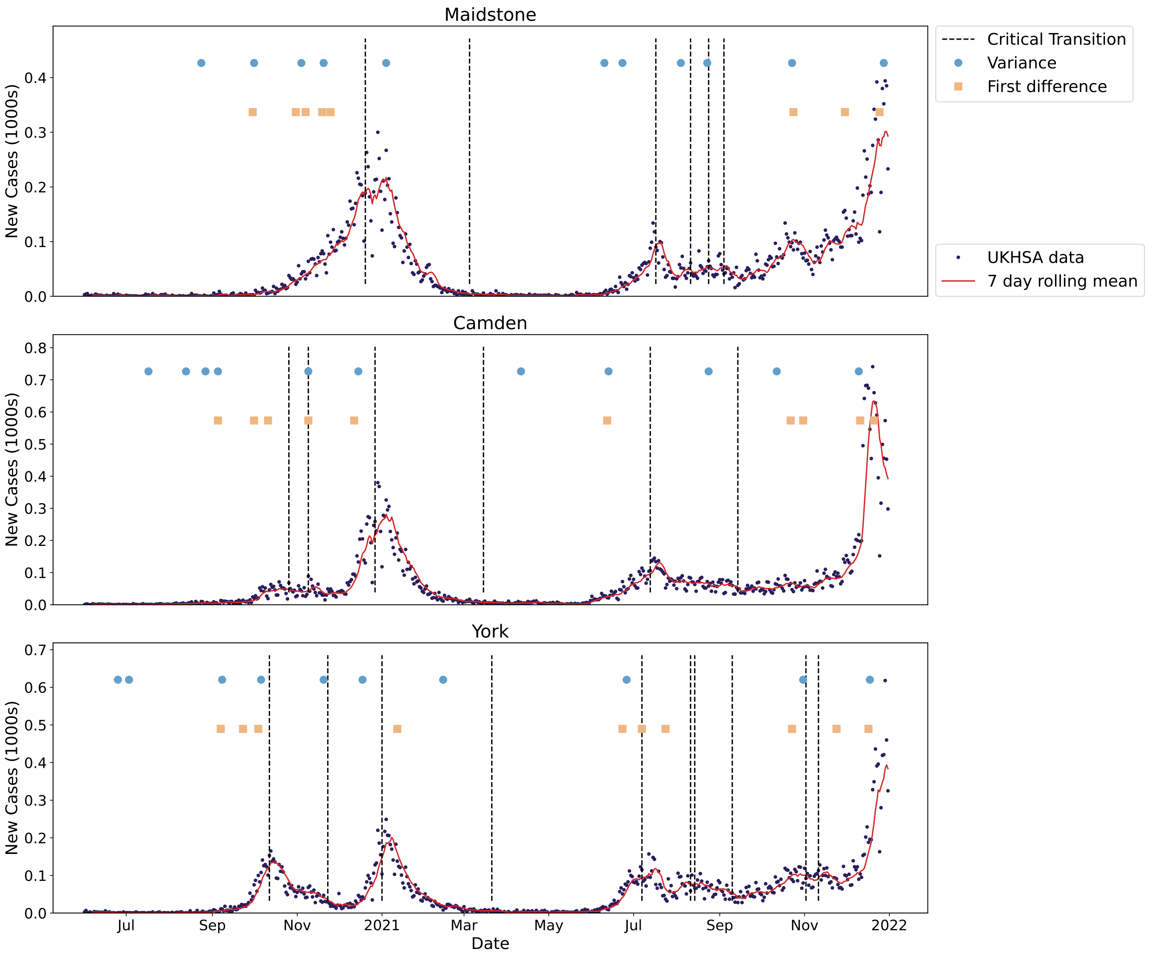


**Fig. S8C.** Illustration of the expected epidemic transitions for a sample of LTLAs between June 2020 and October 2021 alongside the times-of-detection for three consecutive threshold exceedances for the top performing EWS (vertical positioning of the signals does not matter) using a rolling window size of 30. The vertical dotted lines show the expected dates when $R_t = 1$ and the circles and squares show the time-of-detection for variance and first difference in variance EWS. The incidence of new cases and seven-day rolling average are also overlaid.
