## Supplementary material for "Identifying COVID-19 peaks using early warning signals": S9 Figure

### Full NHS results

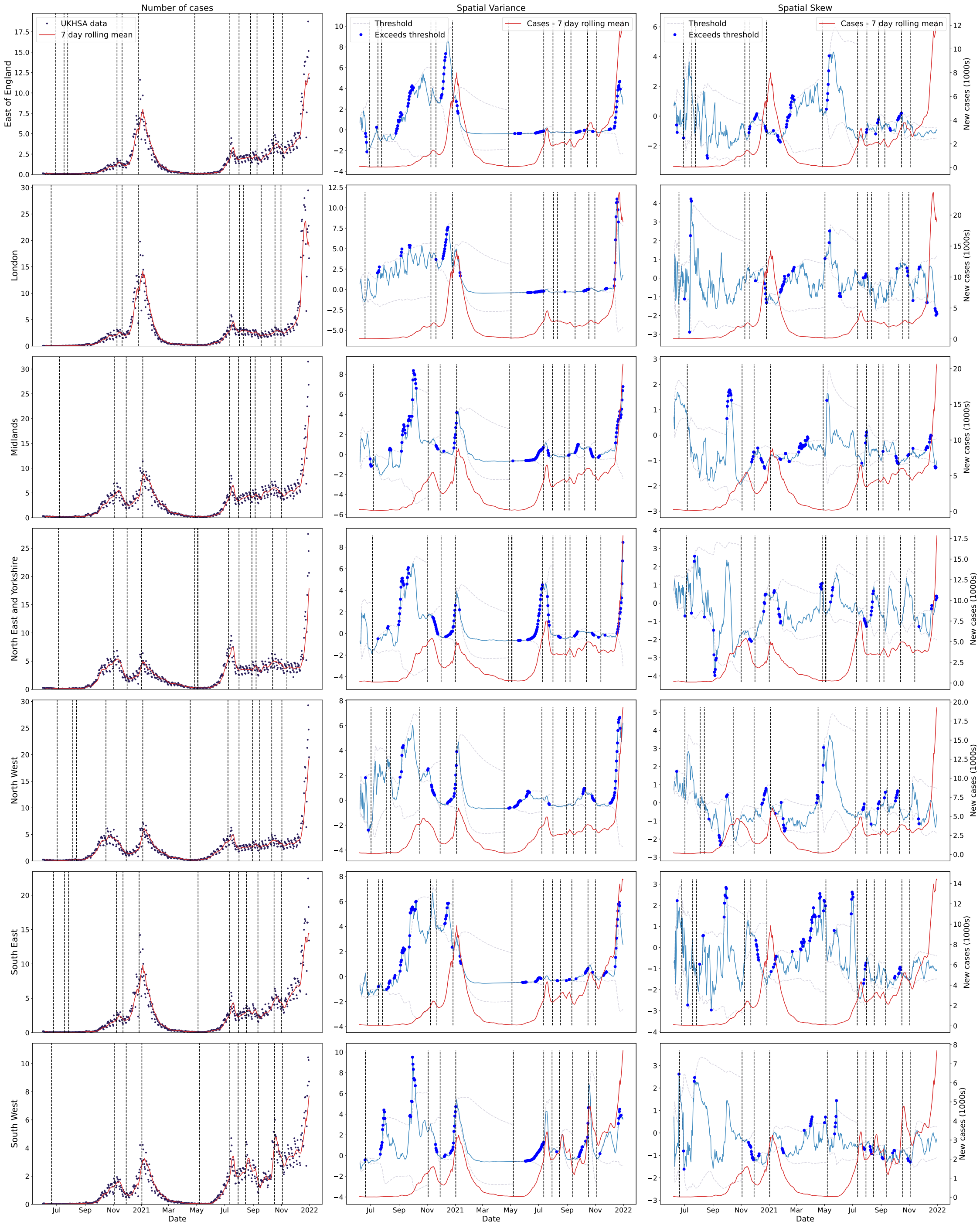


**Fig. S9A.** Incidence and selected, normalised spatial statistics calculated over the spatially detrended reported COVID-19 cases time series data for each of the NHS regions between June 2020 and December 2021. The dotted lines show the 2-$\sigma$ threshold, with dots corresponding to time points where the threshold is exceeded. Incidence data is shown as the dark dots, with the red line being the rolling seven-day average.

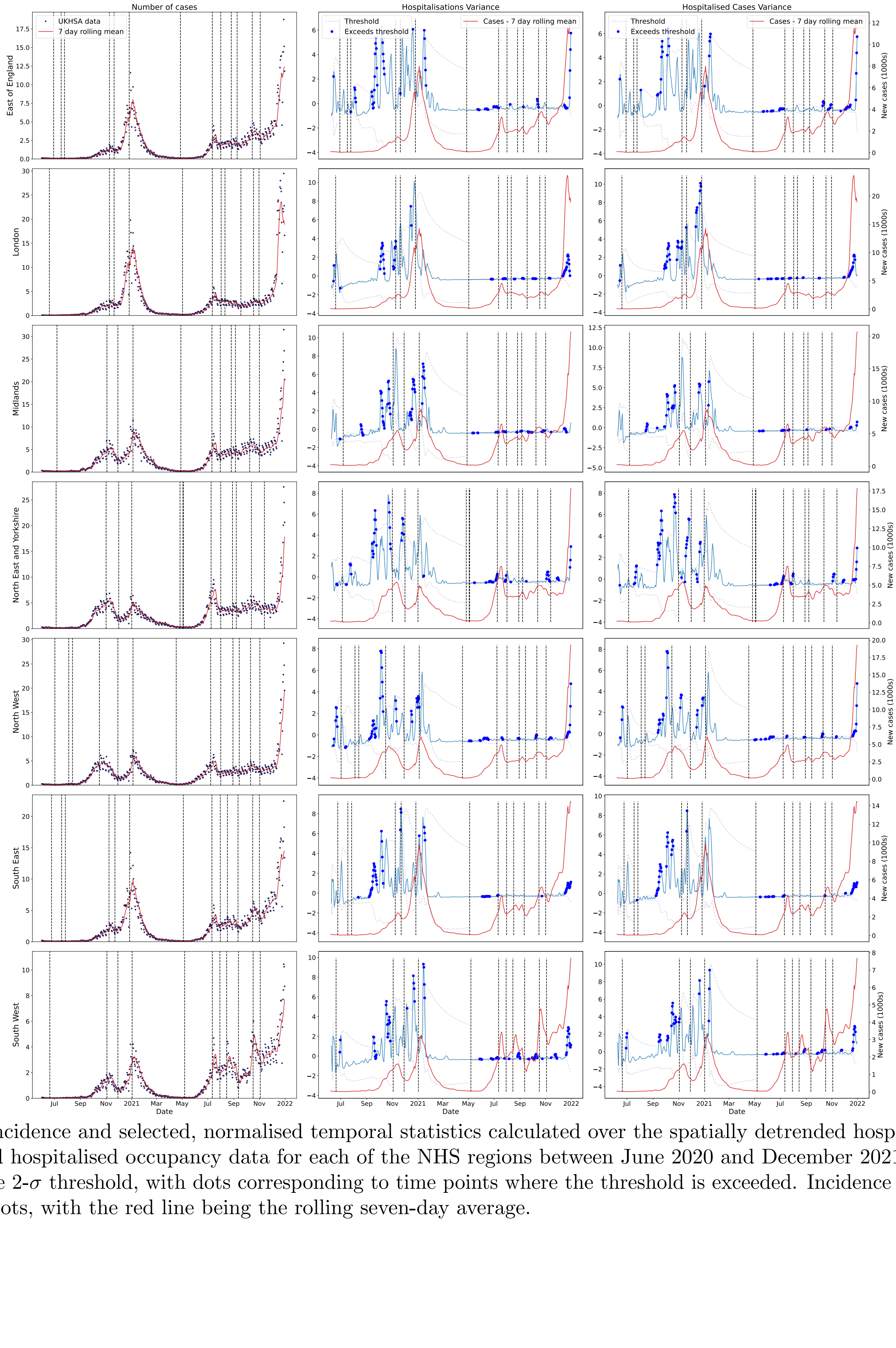


**Fig. S9B.** Incidence and selected, normalised temporal statistics calculated over the spatially detrended hospitalisation incidence and hospitalised occupancy data for each of the NHS regions between June 2020 and December 2021. The dotted lines show the 2-$\sigma$ threshold, with dots corresponding to time points where the threshold is exceeded. Incidence data is shown as the dark dots, with the red line being the rolling seven-day average.
